# The association between socioeconomic status and mobility reductions in the early stage of England’s COVID-19 epidemic

**DOI:** 10.1101/2020.10.28.20221770

**Authors:** Won Do Lee, Matthias Qian, Tim Schwanen

**Affiliations:** Transport Studies Unit, School of Geography and the Environment, University of Oxford, UK; Saïd Business School, University of Oxford, UK

**Keywords:** COVID-19, Everyday mobility, Lockdown, Socioeconomic status, Spatial complexity

## Abstract

This study uses mobile phone data to examine how socioeconomic status was associated with the extent of mobility reduction during the spring 2020 lockdown in England in a manner that considers both potentially confounding effects and spatial dependency and heterogeneity. It shows that socioeconomic status as approximated through income and occupation was strongly correlated with the extent of mobility reduction. It also demonstrates that the specific nature of the association of socioeconomic status with mobility reduction varied markedly across England. Finally, the analysis suggests that the ability to restrict everyday mobility in response to a national lockdown is distributed in a spatially uneven manner, and may need to be considered a luxury or, failing that, a tactic of survival for specific social groups.

## 1. Introduction

Across the planet, government-mandated lockdowns, including restrictions on everyday mobility, are one of the most common policy measures to prevent and reduce the rapid spread of COVID-19 and avoid healthcare services being overwhelmed. While the specific nature of lockdowns varies across and within countries, evidence suggests that they have slowed down the spread of infections (Flaxman et al., 2020; Jarvis et al., 2020; Kraemer et al., 2020; Scally et al., 2020). Lockdowns are, however, drastic interventions with significant economic implications and disproportionally disadvantaging socially and economically vulnerable population groups (Bradbury-Jones and Isham, 2020; Proto and Quintana-Domeque, 2020; Usher et al., 2020). This is why governments in European countries facing a ‘second wave’ of infections hesitate to re-instate nationwide lockdowns.

Not everybody is able or willing to restrict their everyday mobility when lockdowns are in place. Studies using large mobile phone datasets (Oliver et al., 2020; Pepe et al., 2020) have demonstrated significant differences in response to restrictions on everyday mobility. Comparing 65.5m mobile phone GPS traces on 15-17 April with a pre-COVID-19 baseline in the USA, Dasgupta et al. (2020) have found more people staying at home in counties (*n*=2,633) with more healthcare resources, greater wealth and less social deprivation. Bushman et al. (2020) utilised a different dataset with Call Detail Records (CDRs) for ±18m mobile phones across the USA and established that the increase in time spent at home after stay-at-home orders was significantly smaller in Census Block Groups (CBGs) dominated by Blacks, Hispanics and Natives/Other than in those dominated by White and Asians. However, the smaller effect size for the former groups disappeared after income was controlled. The increase was also smaller for CBGs with more individuals aged over 50 than in areas with more younger people, an effect that was independent of income. Finally, a study using spatially more aggregated mobile phone data for 13 regions in France has shown a positive correlation between the prevalence of high standards of living and the percent reduction in mobility after lockdown. It also demonstrates greater mobility reductions in regions with more people aged 24-59, more highly impacted workers and greater numbers of hospitalised people per 100,000 residents (Pullano et al., 2020).

These studies imply that the extent of mobility reduction in an area under lockdown increases as its socioeconomic status (SES) is higher. If true, then this finding can have significant implications for the implementation of future lockdowns and help explain spatial and socioeconomic inequalities in infection, hospitalisation and mortality. For example, the stringency of lockdown rules could be adapted on a local level, taking into account the socioeconomic differences across clinical commissioning groups. The objective of this paper is to provide rigorous proof for the relationship between mobility reduction and SES. Since mobile phone data can be used to monitor people’s response to COVID-19 related restrictions on people’s everyday mobility effectively and on an unprecedented scale (Poom et al., 2020), we use GDPR-compliant CDRs to analyse the relationships between SES and mobility reduction across England while 1) controlling for critical confounding factors and 2) considering spatial complexities in the examined relationships.

England moved to a full government-mandated lockdown later than most other North West European countries. It was only from 23 March 2020 that the Government ordered a reduction of people’s everyday mobility to trips for essential purchases, medical needs and care-giving to others as well as to essential work travel and one stint of exercise per day (Iacobucci, 2020). Gatherings of more than two people who did not live together were prohibited, and all but essential retail premises such as supermarkets were closed. The original restrictions were gradually lifted after 11 May 2020 and have increasingly been replaced by a system of locally specific restrictions (HM Goverment, 2020).

The literature has identified a wide array of factors that are important to mobility patterns. Levels of mobility as represented by distance covered or travel time are conditioned strongly by accessibility levels, which are a combination of the resources (e.g. cars) people have for travel and the configuration of transportation and land-use systems (Fransen et al., 2018; Hanson and Schwab, 1987; Neutens et al., 2010). That configuration can be approximated using the spatial distribution and intensity of people, residences, employment, retail, healthcare facilities, etcetera (Ewing and Cervero, 2010; Newman and Kenworthy, 1996). Everyday mobility is also shaped by commitments on people’s time use demanding they are at certain places in physical space at particular times are also shaping everyday mobility (Cullen and Godson, 1975; Schwanen et al., 2008; Van Acker et al., 2010). The health of people in a given area should also be controlled when the relationship between SES and reduction in mobility is analysed. This is because COVID-19 tends to pose greater risks to people with underlying health conditions (Fletcher et al., 2020; Jordan et al., 2020), and such conditions are likely to be correlated with both SES and mobility levels (O’ Lenick et al., 2017; Yoo et al., 2018). It is also vital to consider SES as a multi-dimensional construct in the analysis of spatial variation in everyday mobility (Hanson and Hanson, 1981; van de Coevering and Schwanen, 2006; Xu et al., 2018). This is why the analysis considers not only household income but also education and skills, occupation level, housing tenure and crime levels in residential areas. In short, this paper analyses the relationships between multiple indicators of SES and the reduction in everyday mobility during the spring 2020 lockdown in England, while controlling for the effects of accessibility, activity commitments and population health.

Relevant spatial complexities include spatial dependencies and spatial heterogeneity (Anselin, 1988). The former follows from Tobler’s (1970) claim that geographical phenomena that are near to each other are often related to each other, and which still resonates in the current era of unprecedented global interconnections (Miller, 2004). Spatial dependencies manifest as spatial autocorrelations in linear models, potentially resulting in inaccurate conclusions about the associations between dependent and independent variables. This risk is particularly salient when spatial dependencies occur in the dependent variable (as correlations in the levels of mobility reduction in adjacent or near areas in our case) or the residuals (as correlations in the values of unobserved variables in adjacent or near areas). Spatial heterogeneity refers in the current context to geographical scale: the relationships between SES and mobility reduction may not be the same across all parts of the study area (England) but exhibit regional variations (Bonaccorsi et al., 2020; Scala et al., 2020). This suggests that the national level is not necessarily the appropriate scale at which to evaluate the relationship. Both spatial dependencies and spatial heterogeneity will be considered below as various studies have suggested they shape the relationships of demographic, environmental and healthcare factors with COVID-19 infection, hospitalisation and mortality across regions (Harris, 2020; Mollalo et al., 2020; Perone, 2021; Xiong et al., 2020).

## 2. Data and methods

### 2.1 Data and variables

The analysis used anonymised and aggregated GDPR-compliant CDRs for 1,119,449 users in the period 1 March-18 April 2020^1^. CKDelta, a company that collected, cleaned, and anonymized the mobile phone location data from a large British mobile phone provider, granted us access to the dataset. An entry is created within the CDRs based on the on-the-fly processing of signalling messages exchanged between mobile phones and the mobile network, usually collected by mobile network operators to monitor and optimise the mobile network activities. CDRs encompass messages containing information about the identifiers of the user and of the cell phone tower handling the communication and the time stamp.

The temporal resolution of the CDRs, reflected in the timestamps of the user’s activity, is at a second level. The median user has 42 daily CDR records with corresponding location observations before the lockdown and 24 records afterward. The spatial resolution is on a cell phone tower level. Within urban areas, the average distance from one cell phone tower to its closest neighbour is around 300 meters. In rural areas, the cell phone tower density is much lower, and cell towers are one to two kilometres apart, with the largest distance to the nearest neighbouring tower at 12 kilometres.

We use CDRs to compute the daily median radius of gyration for users in each Clinical Commissioning Group (CCG) area (*n*=191), which is a generic metric of the spatial extent of everyday mobility practices for a user. We compute the median instead of the sample mean of the radius of gyration for individual users to avoid the upward bias from the right tail of the distribution who represent those users travelling long distances on a given day. Thus, our median measure is unaffected by shifts in the long-range displacements of users. In general, the radius of gyration is a function of the number of places a user visits on a day and the distance between those places. In most cases, the radius of gyration increases if the user visits more places or if the distance between the visited places is larger. The radius of gyration is unaffected by the amount of time users spend away from their residence. Mobility will be underreported if individuals do not take their phone along or when there are no CDR records generated during a trip.

Alternative measures of both the range and amount of mobility exist. Pullano et al. (2020) consider long trips, which are identified with cell phone towers being more than 100 km apart. Qian et al. (2020) consider the flow of users between clinical commissioning groups and the share of users staying at home.^2^ Hoteit et al. (2014) also discuss the total trajectory length of the users as a direct measure of the amount of mobility. However, they note that the range and amount of mobility are highly corrected. While other measures of mobility have equal merit, we use the radius of gyration as it is widely employed in studies using mobile phone data (González et al., 2008; Pappalardo et al., 2019, 2015).

Researchers have provided various interpretations for our gyration measure. González et al. (2008) interpreted the measure as the characteristic distance travelled by a user. De Montjoye et al. (2013) explained it as the smallest circle that contains all the places a user has been to on a given day. Xu et al. (2018) suggested that it measures the spatial extent of a phone user’s activity space from the user’s trajectories in a given time. To this end, this research measures the extensivity of activity space (Lu et al., 2020) though summation of the distance from all points of user *i* travels among the time-stamped (*t*) locations *l*_*i,d,t*_ on day *d* from the trajectory’s centre of mass *c* can be formulated as 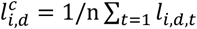 on that day. Locations *l* are approximated by the nearest mobile phone tower. Formally, the radius of gyration r_i,d_ can be expressed as:

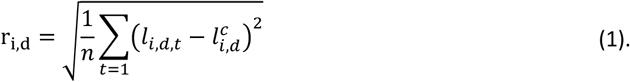

We aggregate user-level r_i,d_ values to the spatial level CCG areas in which individuals reside,^3^ to protect the individual privacy and because CCGs decide what healthcare services are needed to meet the specific needs of the population in their area, and make sure those services are provided. While NHS (National Health Service) England hold primary responsibility for the commissioning of primary care services such as general practitioner (GP) and dental care services, CCGs commission most hospital care, including urgent and emergency care. They thus play a critical role in the healthcare response to COVID-19; their services will be overwhelmed first if compliance with a government-mandated lockdown is (very) low. We use the median r_i,d_ across users in a given CCG area in the analysis below, because the median is less sensitive to right-skewed distributions and outliers than the sample mean. Because the study concentrates on mobility reductions, it does not analyse the median radius of gyration of residents across CCG areas *j* on a given day, *G*_*j,d*_ It rather analyses the reduction in mobility *R*_*j,d*_ relative to a reference day, *REF:*

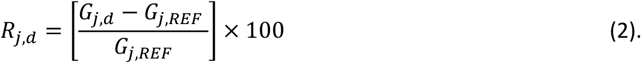

Tuesday 3 March is chosen as *REF*, because it was the last ‘normal’ Tuesday before mobility levels in England started to drop in connection with COVID-19. Prior to the epidemic Tuesday was the day of the week on which mobility levels tended to be very high because the share of people commuting to employment or education was larger than on most other days (Department for Transport, 2020a). As explained in Section 3.1, we study population movement on four consecutive Tuesdays starting on the 24 March, which is the day after the national lockdown commenced, until the 14 April.

The information on mobility reductions across England is correlated with data on SES, accessibility, activity commitments and population health from the 2011 Census and other sources provided by the Office for National Statistics (ONS). While a Lower Layer Super Output Area (LSOA) level is the spatial resolution of non-mobility datasets, we aggregate data to the CCG area level by using an official lookup table retrieved from ONS geography portal. The creation of variables from the available data has been based on the authors’ knowledge of relevant literature on the spatial variation in everyday mobility (see Introduction). In total, we use 28 operational variables in the regression analyses described below (Table 1). Originally, a series of indicators of the racial/ethnic composition of the CCG areas were included but these had to be dropped because of multicollinearity with the percentage of people who do not speak English and resident population density. Spatially disaggregated data on COVID-19 incidence rates were also excluded from the analysis because of low reliability in March and April 2020 when diagnostic testing for the disease was very limited in England.

**Table 1.**
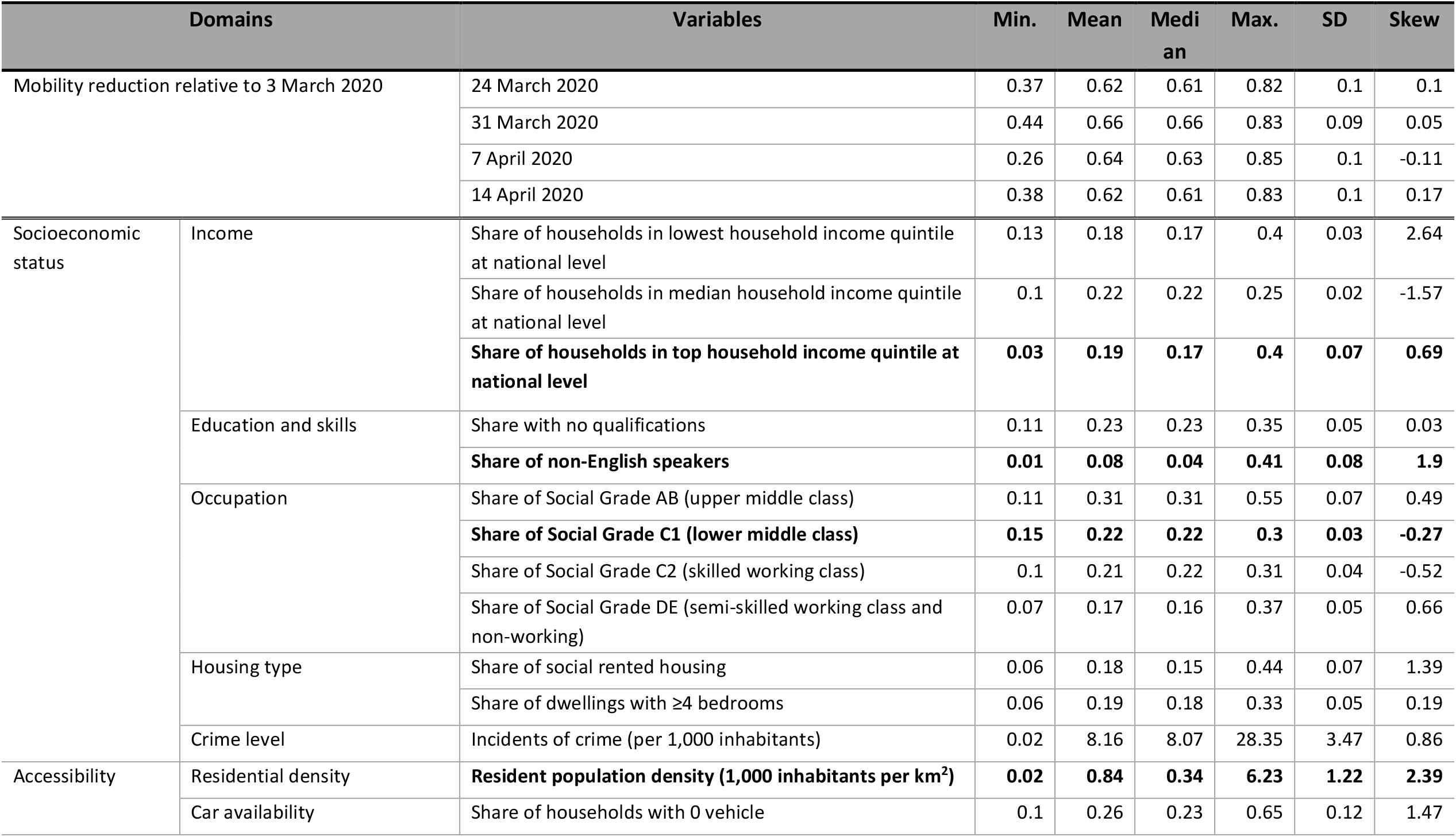
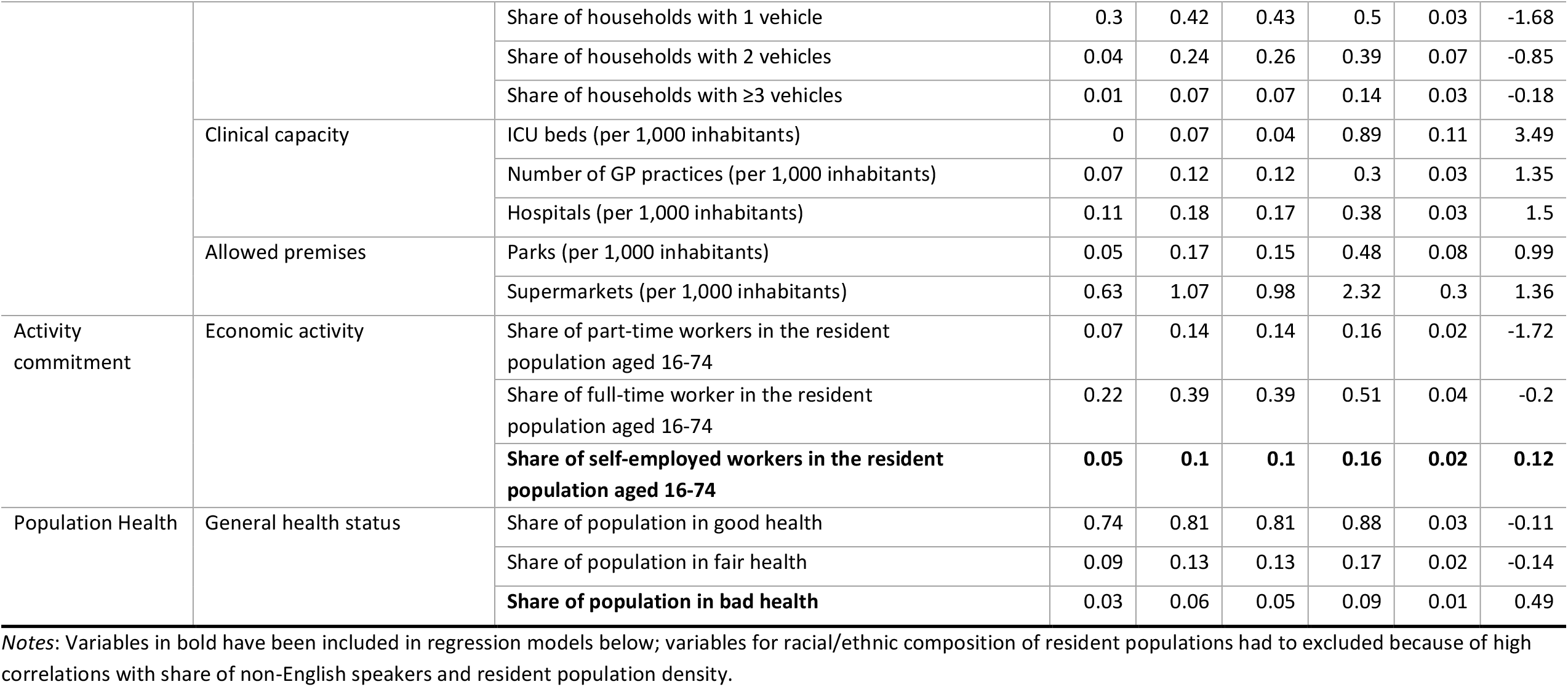
Descriptive statistics of variables.

### 2.2 Modelling

A series of econometric models of increasing spatial complexity has been specified to understand the relationships of SES, accessibility, activity commitments and population health with mobility reduction *R*_*j,d*_. The first model is a Stepwise Linear Regression Model (SLRM) in which the values for the dependent variable and regressors are treated as independent from each other is given by *R*_*j,d*_= β_0_ + *x*_*j*_β_*j*_+ *ε*_*j*_, where β_0_ is the intercept, *x* a vector of regressors, β a vector of regression coefficients, and *ε* the error term. We used a combination of forward and backward elimination of potential explanatory variables to achieve a specification that combines model parsimony with goodness-of-fit and plausibility of interpretation. All variables have been standardised with a mean of zero and variance of unity to facilitate interpretation (Oshan et al., 2020). The contribution of each regressor in the SLRM to the statistical explanation of mobility reduction is demonstrated with the help of Lindeman et al.’s (1980) relative importance metric (*LMG*), which identifies the average incremental improvement of each predictor based upon the decomposition of *R*^2^.

Next, we consider two distinct spatial regression models that account for spatial autocorrelation at the global level of all 191 CCG areas in England, as defined by spatial weights *W* (Anselin and Arribas-Bel, 2013). The Spatial Lag Model (SLM) captures the substantial spatial dependency in mobility reduction in a given CCG area and the neighbouring CCGs. The SLM model specification includes a spatially lagged dependent variable:

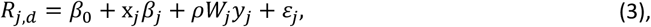

where the spatial lag coefficient *ρ* indicates the impact of mobility reductions in neighbouring areas on the reduction in CCG area *j*. In contrast, the Spatial Error Model (SEM) addresses spatial error autocorrelation. The SEM model specification includes a spatial autoregressive error term:

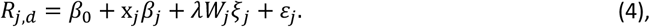

where *ξ*_*j*_ denotes the spatial component of the error term and *λ* the spatial error coefficient.

Finally, we use two different Geographically-Weighted (GW) models to examine the local variation in the rates of change so that the coefficients in the model are specific to a location *u*_*j*_, *v*_*j*_ rather than being global estimates. Geographically Weighted Regression (GWR) estimates the *k*th coefficient β_*k*_ for location *u*_*j*_, *v*_*j*_ through kernel density estimation (Fotheringham et al., 2017). The GWR regression specification can be expressed as:

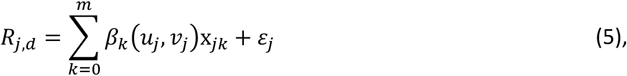

Where *m* = *n* + 1 (191 CCG areas+1=192). To obtain the localised parameter estimates β_*k*_ *u*_*j*_, *v*_*j*_ for each regressor x_*jk*_, GWR employs the (diagonal) spatial weight matrix *W*, constructed from the weights to define the spatial neighbourhood that provides the best model fit. The kernel density estimation approach requires the specification of a kernel function and a bandwidth for *W* (Brunsdon et al., 2010). This study uses the bi-square kernel function and calibrates the bandwidth on the basis of *k*=4 nearest neighbours to generate the local weightings (Li et al., 2020; Oshan et al., 2019). It also uses a fixed average bandwidth, *BW*, of 150 across all regressors. The vector of local parameters 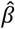 for the matrix of regressors *X* can be expressed as:

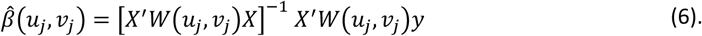

While the GWR captures spatial heterogeneity in a manner that the global models SLRM, SLM and SEM cannot do, it assumes that the spatial scale over which local variations in the effects of regressors – and thus the spatial processes they reflect – are identical across regressors. This assumption may be unduly limiting and is relaxed in Multi-scale Geographically Weighted Regression (MGWR). MGWR allows for conditional relationships between the dependent variable and each regressor through the selection of an optimal BW for each regressor (Fotheringham et al., 2017) can be expressed as Equation (7):

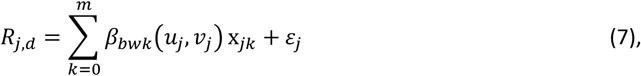

where *bwk* indicates the BW used for calibration of the *k*th conditional relationship for MGWR. This study also deploys the adaptive kernel with bi-square function for MGWR model calibration to update and remove the spatial effects of each regressor outside the neighbourhood as specified by bandwidth. All GW models in this study were estimated using the *GWmodel* package in *R* software (Gollini et al., 2015).

## 3. Results

### 3.1 Spatial and temporal patterning of mobility reduction in England

Across England, mobility levels as represented by the median radius of gyration declined 70.4% between 3 and 28 March (Qian et al., 2020; Santana et al., 2020). This reduction commenced almost two weeks before the start of the government-mandated lockdown on 23 March, and over the course of April gradually reversed. Nonetheless, the England-wide trend in Figure 1 disguises stark local and regional variations. Figure 2 depicts hot and cold spots in mobility reduction based on the *Gi*\* statistic, which identifies local pockets where particular values for a given variable are concentrated (Getis and Ord, 1992; Ord and Getis, 1995). Hot and cold spots are spatial clusters of contiguous CCG areas where mobility reductions are significantly (at *p*<0.05) greater or smaller than the England-level average. Figure 2 shows multiple spatial clusters, with the most considerable decrease in everyday mobility (i.e. hot spots) occurring in Greater London and the smallest (i.e. cold spots) in the Yorkshire and the Humber region and South West England. The locations of hot and cold spots bear some resemblance to the hotspots of COVID-19 incidence rates during spring 2020 in England (NHS Digital, 2020). According to the Royal Berkshire NHS Trust, London recorded the first COVID-19 related death on the 5 March. Yorkshire and the Humber and South West England recorded their first deaths only on 17 March and 15 March, respectively.

**Figure 1.**
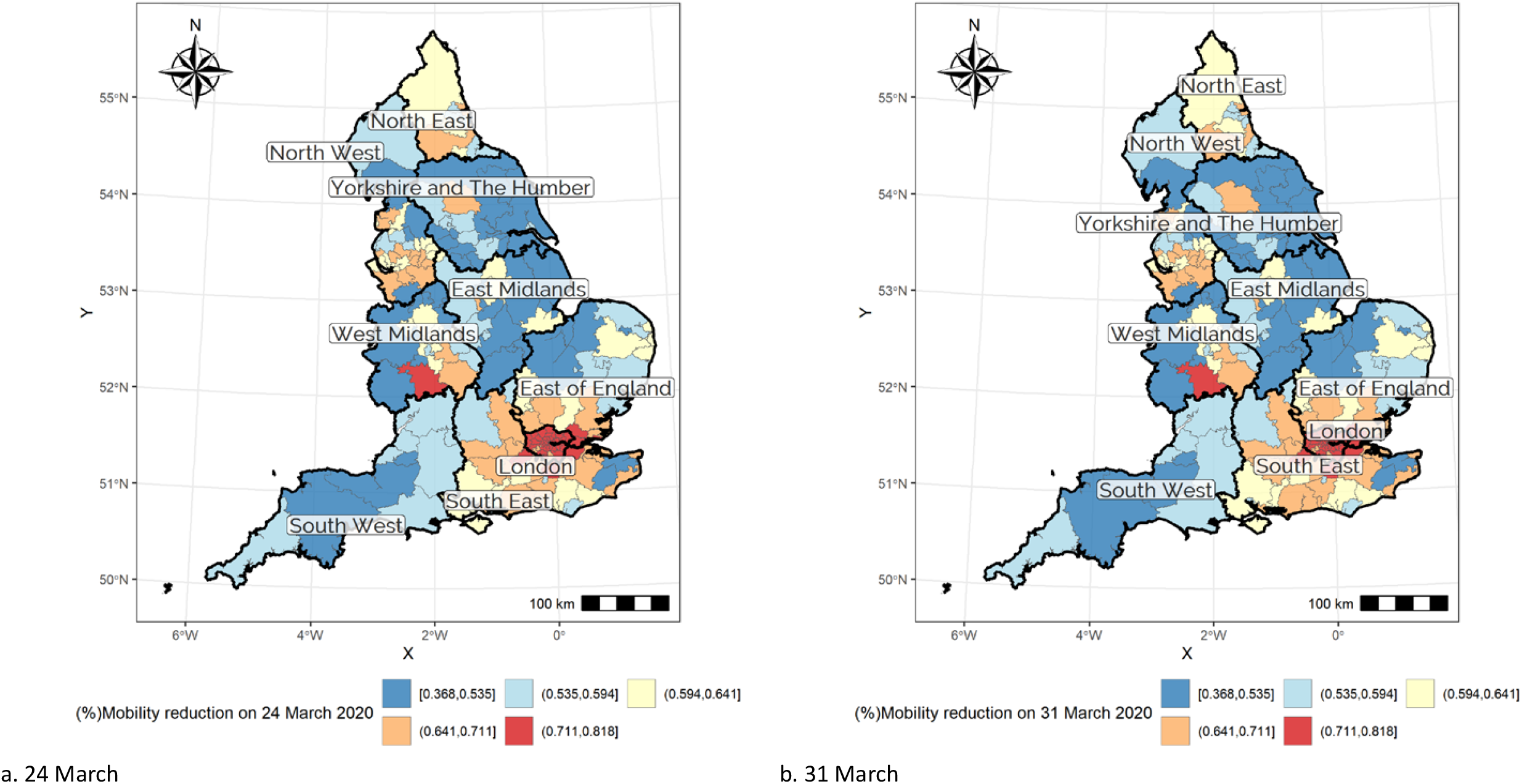
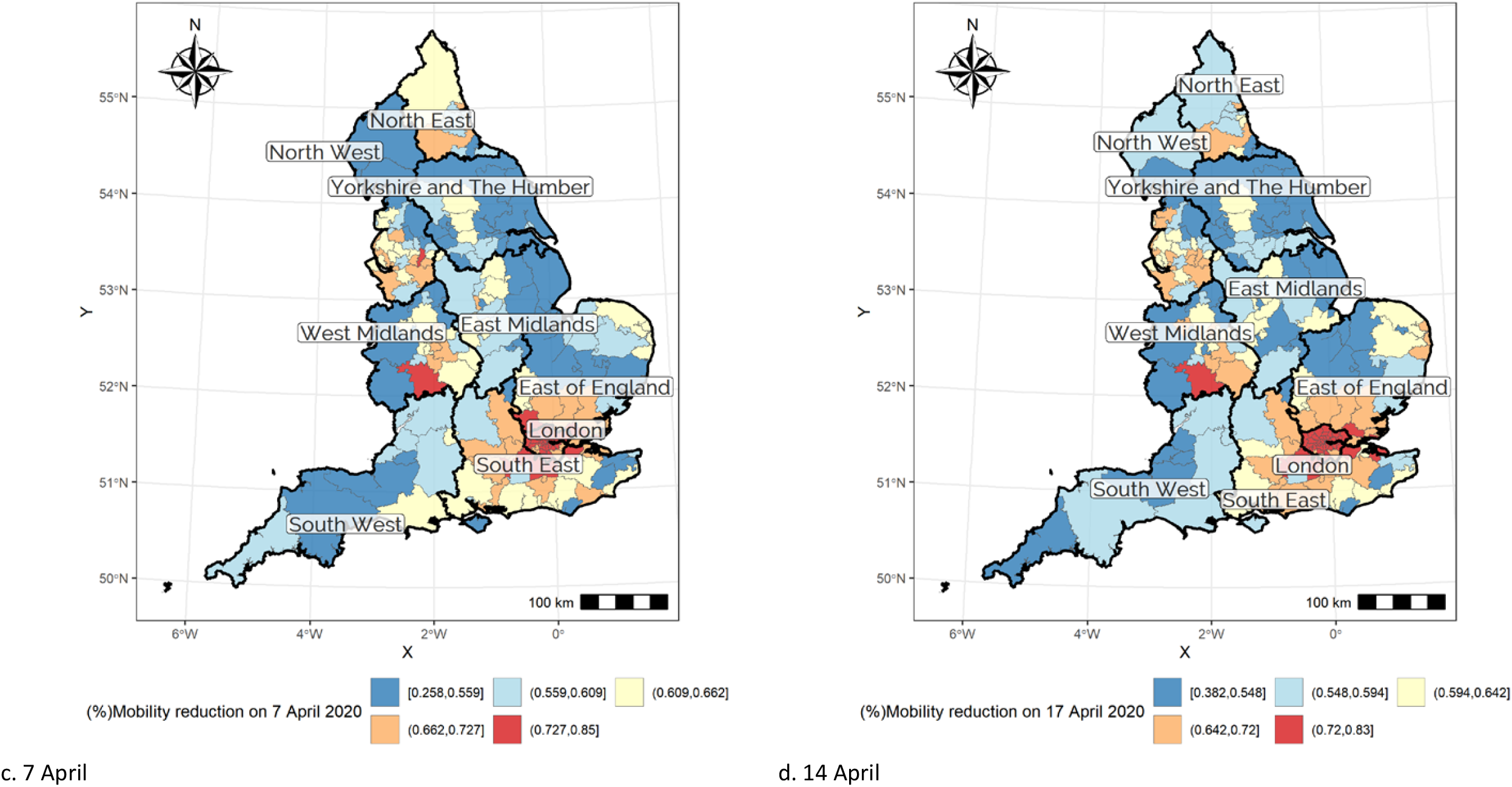
Maps of mobility reduction across England, by date.

**Figure 2.**
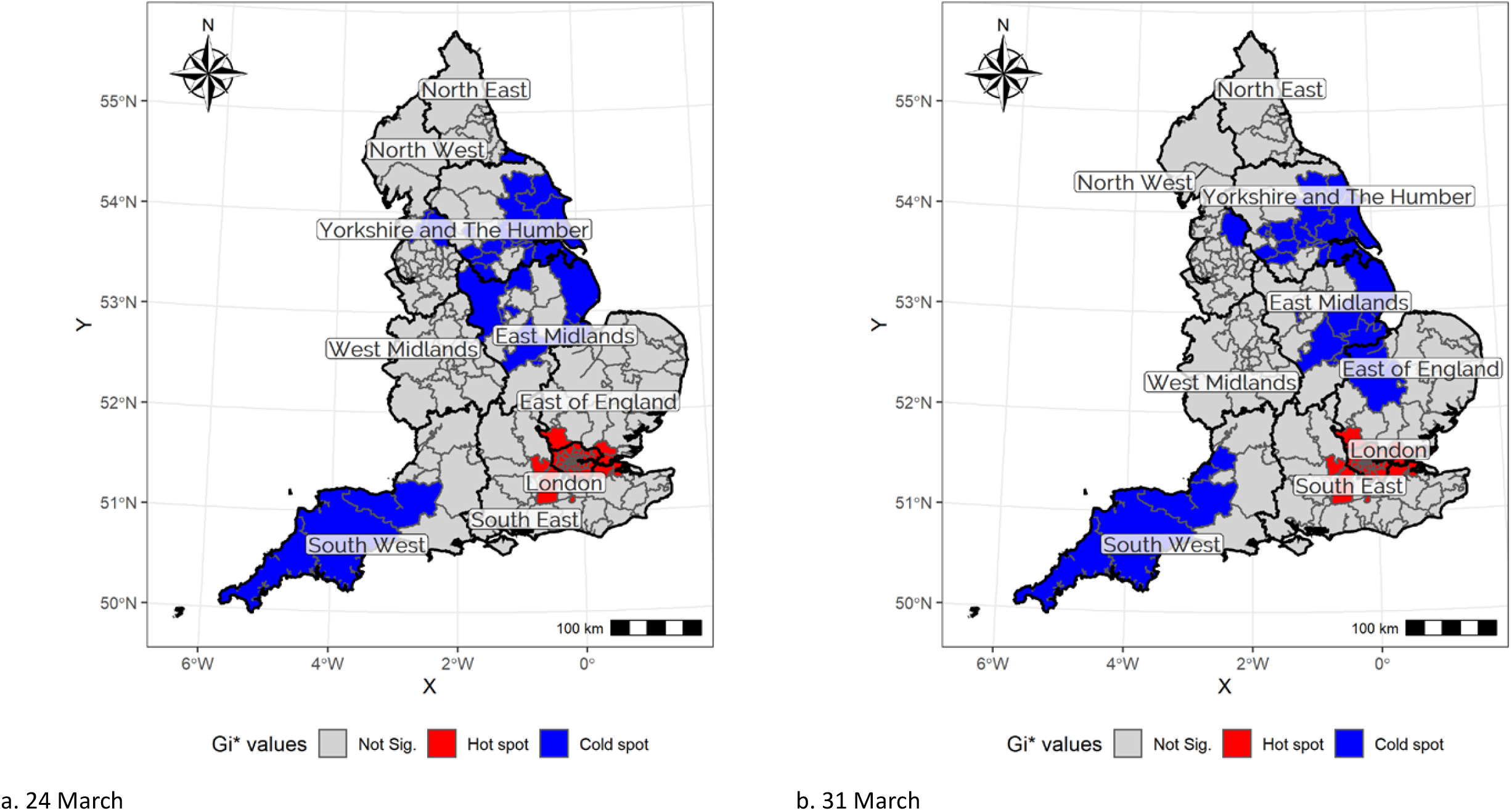
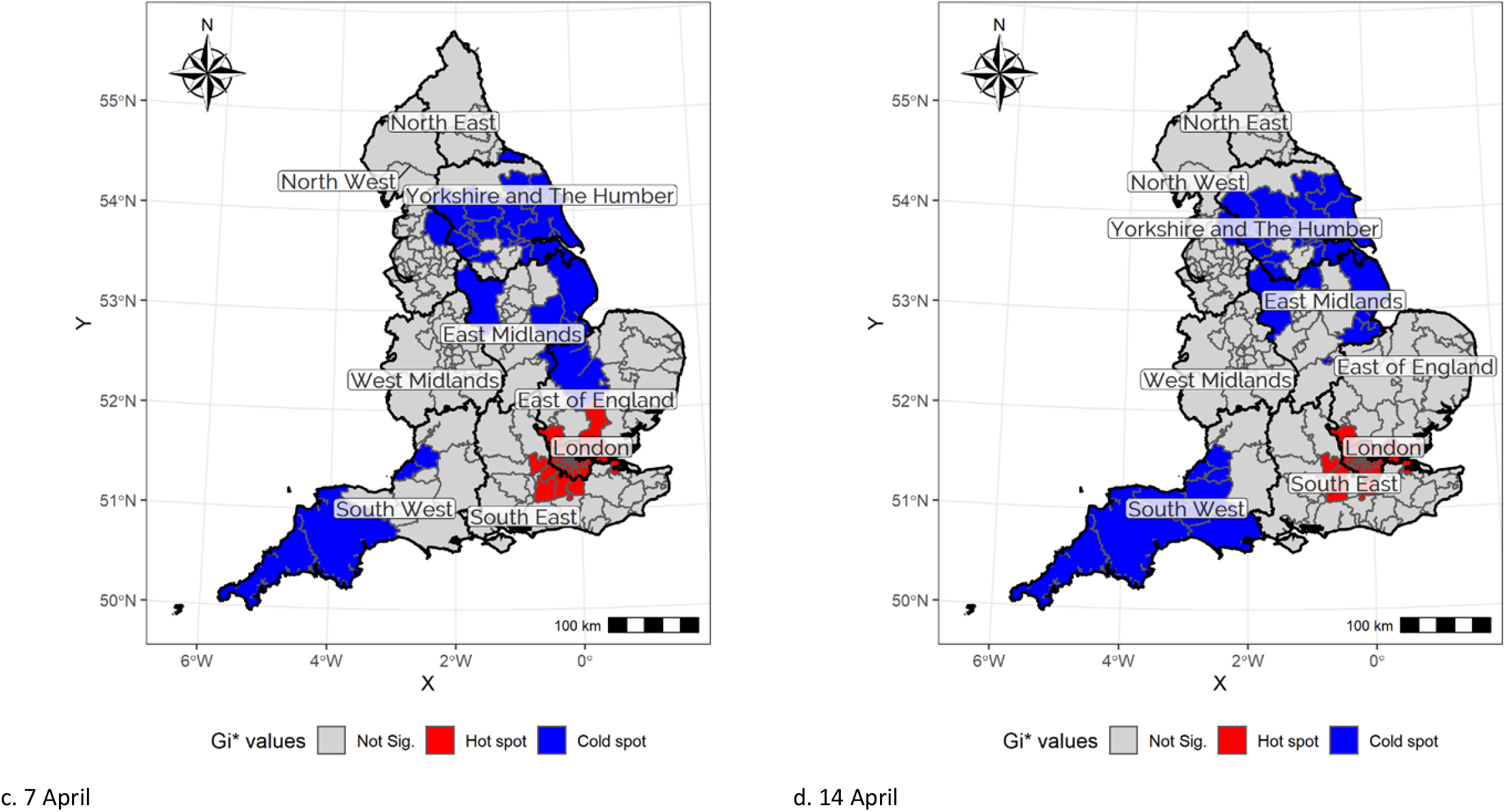
Maps of hot and cold spots in mobility reduction across England, by date.

### 3.2 The association of socioeconomic status with everyday mobility reductions

To some extent, and across all four dates considered, the hot and cold spots of mobility reduction map onto areas where households in respectively the top and bottom quintiles of the national income distribution are overrepresented. Greater London clearly has the most households in the top income group, and Yorkshire and the Humber has many in the lowest quintile (Figure 3). A visual comparison of Figure 2 and 3 suggests a correlation between the reduction in everyday mobility levels and SES of CCG areas. It is, however, unclear how strong and linear the association with income, as only one marker of SES, is and whether other factors confound that association.

**Figure 3.**
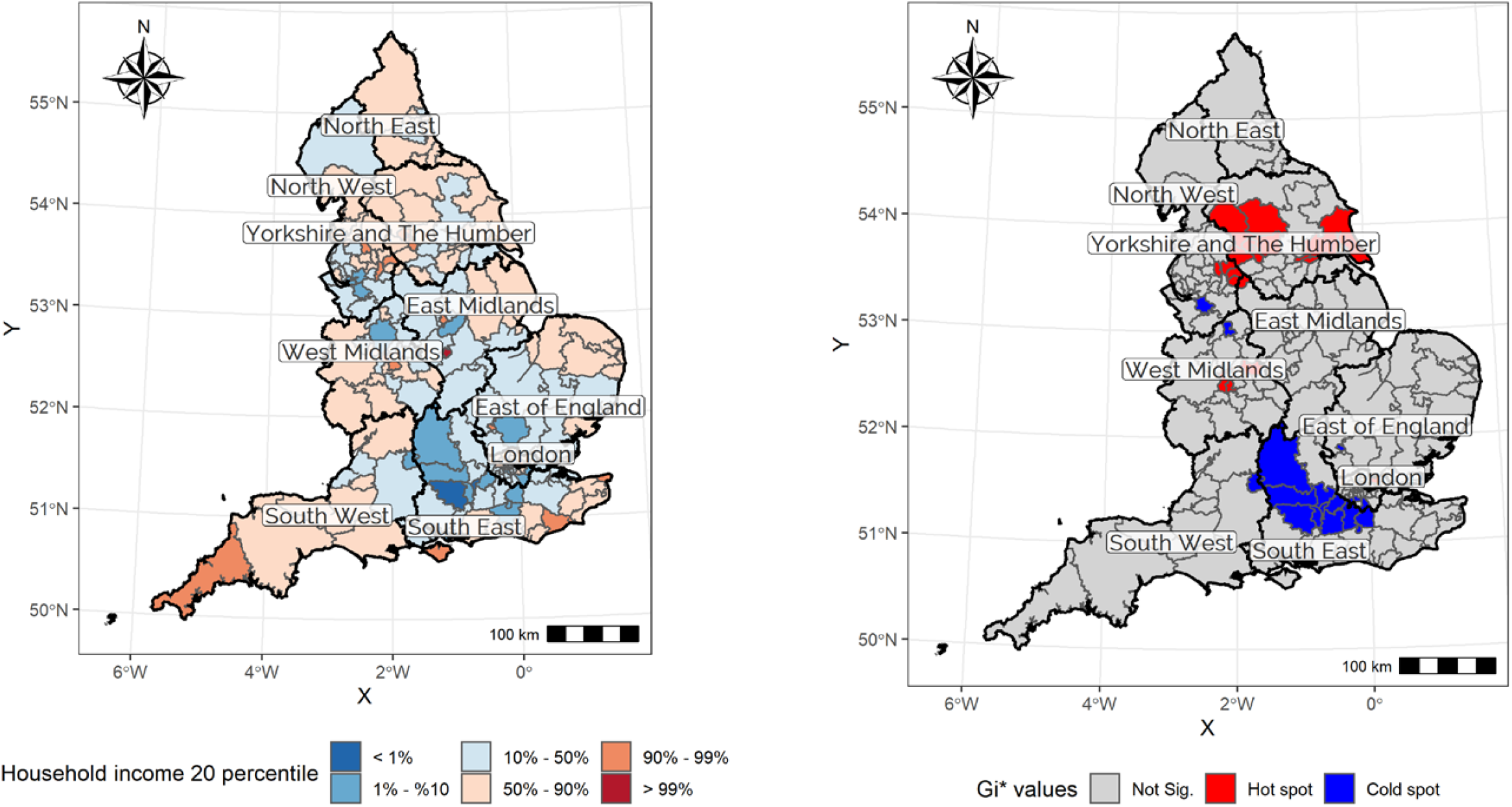
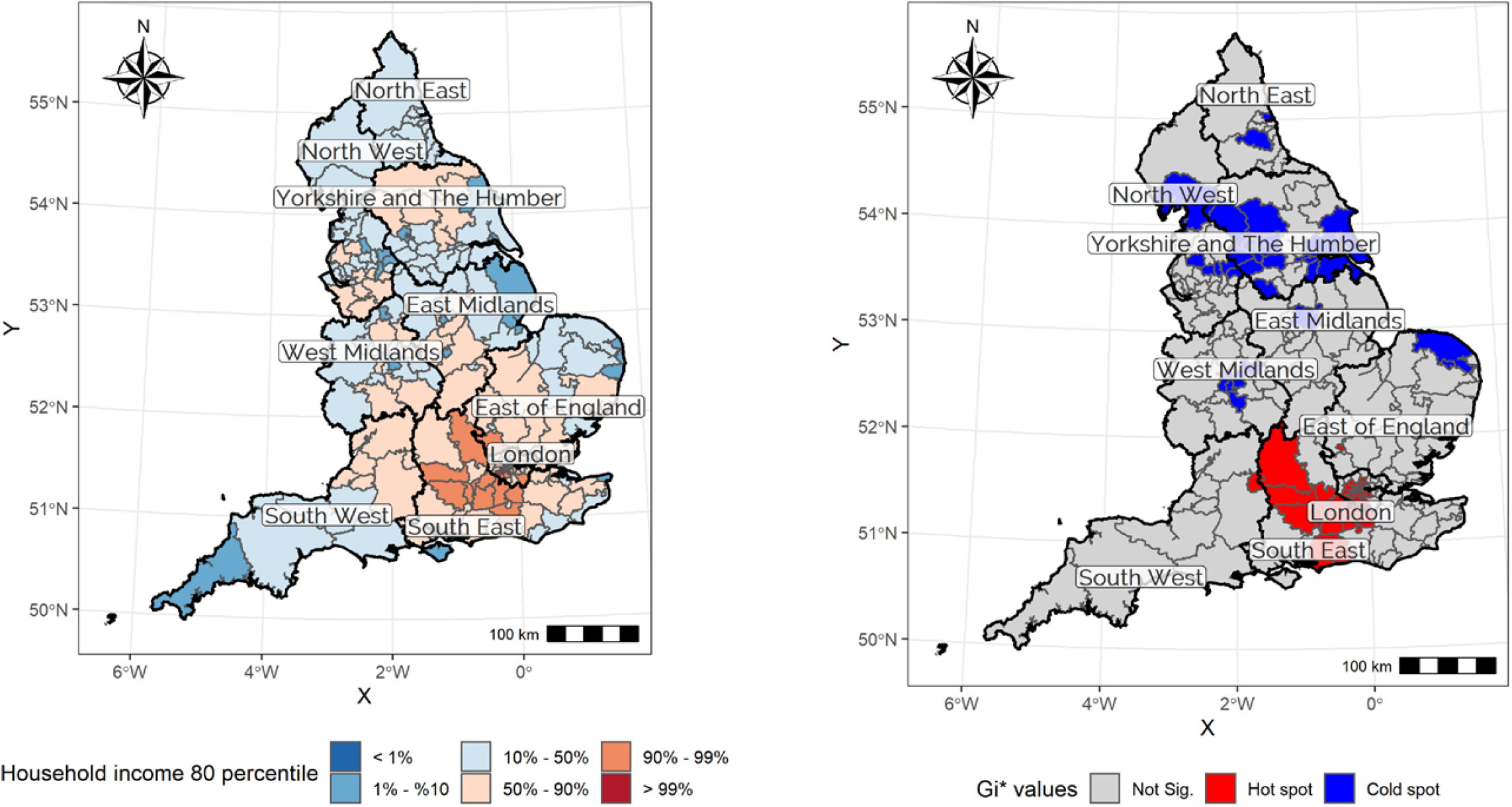
Spatial distribution of the lowest (above) and highest quintiles (below) of household income across England.

The Stepwise Linear Regression Model (SLRM) confirms a clear relationship of income with mobility reduction and demonstrates that multiple other variables help to explain spatial differences in mobility reduction during the government-mandated lockdown across England. Of the 28 variables considered, only six are significantly (*p*<0.01) correlated with the level of mobility reduction that multicollinearity does not affect (*VIF*<6). The selected variables are identical across the four consecutive Tuesdays in March and April. Half relate to SES of the resident population: share of non-English speakers, share of high-income households who belong to the top quintile of the household income distribution in England and Wales, and share in lower-middle class occupation (social grade C1). The other three selected variables concern accessibility (resident population density), activity commitments (share of population who are self-employed), and population health (share of residents with bad health status). Depending on the date considered, these six variables explain 70-74% of the variation in the spatial distribution of mobility reduction.

The regression coefficients show that mobility reduction was greater in areas with more high-income households, non-English speakers, workers in lower middle-class occupations and people in bad health. Mobility reduction was also greater in areas with higher average population density but smaller in those with more self-employed workers. The effect sizes are fairly stable across the four dates considered, and sensitivity tests have shown that all relationships are approximately linear in nature. According to the *LMG* values, the household income variable makes the strongest contribution to the explanation of spatial differences in mobility reduction, followed by residential density, then the share of people who do not speak English. A local area’s SES was strongly associated with the extent of the mobility reduction among its residents at the start of the national lockdown.

### 3.3 Spatial complexity I: The importance of spatial dependency

While useful, SLRM models are spatially naïve because they do not account for spatial dependency or heterogeneity. The spatial clustering of mobility reduction levels across England in Figure 2 gives reason to expect unobserved factors may be creating spatial dependencies that potentially bias the regression coefficients. The SLM models, in which a spatially lagged dependent variable – i.e. the average mobility reduction in surrounding areas – is included as an additional regressor, confirms the existence of upward bias in the standard regression model. Table 2 show a significant spatial lag coefficient on all four dates, yet it also displays smaller coefficients for the six previously selected variables. The Akaike Information Criterion (AIC), which measures the extent of information loss for a given model, can be used to compare different model specifications with each other (as long as the more restricted version is nested in the other). The smallest difference Δ*AIC*_*SRM*_ occurs for 31 March with a value (2.0) that still offers, in the words of Burnham and Anderson (2004, page 70), “substantial” evidence in favour of the SLM specification; for the other dates the evidence favouring the SLM specification is considerably stronger.

**Table 2.** Spatial regression modelling of mobility reduction, by date.

|  | 24 March 2020 |  | 31 March 2020 |  | 7 April 2020 |  | 14 April 2020 |  |
| --- | --- | --- | --- | --- | --- | --- | --- | --- |
|  | SLM | SEM | SLM | SEM | SLM | SEM | SLM | SEM |
|  | Coeff (SE) | Coeff (SE) | Coeff (SE) | Coeff (SE) | Coeff (SE) | Coeff (SE) | Coeff (SE) | Coeff (SE) |
| **(Intercept)** | -0.009<br>(0.037) | 0<br>(0.041) | -0.008<br>(0.038) | 0<br>(0.04) | -0.013<br>(0.037) | 0.002<br>(0.053) | -0.012<br>(0.035) | -0.001<br>(0.045) |
| ***Socio-economic status*** |  |  |  |  |  |  |  |  |
| Share of households in top household income quintile at national level | 0.666***<br>(0.083) | 0.795***<br>(0.073) | 0.7***<br>(0.086) | 0.806***<br>(0.073) | 0.612***<br>(0.084) | 0.794***<br>(0.082) | 0.636***<br>(0.08) | 0.764***<br>(0.074) |
| Share of non-English speaker | 0.287***<br>(0.073) | 0.365***<br>(0.071) | 0.336***<br>(0.076) | 0.399***<br>(0.072) | 0.248***<br>(0.074) | 0.374***<br>(0.078) | 0.232***<br>(0.069) | 0.337***<br>(0.071) |
| Share of social grade C1 (lower middle class) | 0.428***<br>(0.062) | 0.492***<br>(0.062) | 0.475***<br>(0.065) | 0.535***<br>(0.062) | 0.394***<br>(0.063) | 0.443***<br>(0.068) | 0.429***<br>(0.06) | 0.486***<br>(0.062) |
| ***Accessibility*** |  |  |  |  |  |  |  |  |
| Resident population density (1,000 inhabitants per km<sup>2</sup>) | 0.284***<br>(0.08) | 0.353***<br>(0.081) | 0.284***<br>(0.082) | 0.341***<br>(0.082) | 0.267***<br>(0.081) | 0.31***<br>(0.088) | 0.306***<br>(0.076) | 0.363***<br>(0.081) |
| ***Activity commitments*** |  |  |  |  |  |  |  |  |
| Share of self-employed workers | -0.186***<br>(0.06) | -0.196***<br>(0.063) | -0.229***<br>(0.062) | -0.245***<br>(0.064) | -0.24***<br>(0.061) | -0.238***<br>(0.069) | -0.2***<br>(0.057) | -0.2***<br>(0.063) |
| ***Population health*** |  |  |  |  |  |  |  |  |
| Share of population in poor health | 0.397***<br>(0.075) | 0.465***<br>(0.074) | 0.435***<br>(0.078) | 0.492***<br>(0.075) | 0.335***<br>(0.076) | 0.446***<br>(0.082) | 0.356***<br>(0.072) | 0.441***<br>(0.075) |
| ***Spatial autoregressive parameters*** |  |  |  |  |  |  |  |  |
| Spatial lag coefficient ($\rho$) | 0.22**<br>(0.076) | - | 0.178**<br>(0.078) | - | 0.31***<br>(0.073) | - | 0.272***<br>(0.072) | - |
| Spatial error coefficient ($\lambda$) | - | 0.077<br>(0.106) | - | 0.039<br>(0.109) | - | 0.28***<br>(0.094) | - | 0.206*<br>(0.099) |
| ***Model fit metrics*** |  |  |  |  |  |  |  |  |
| Log-likelihood | -143.0 | -145.7 | -148.7 | -150.8 | -146.7 | -150.6 | -134.3 | -138.7 |
| Pseudo $R^2$ | 0.738 | 0.729 | 0.721 | 0.714 | 0.726 | 0.715 | 0.76 | 0.749 |
| $AIC_{SLRM}$ | 307.9 | | 317.8 | | 323.5 | | 296.6 | |
| $AIC_{SRM}$ | 303 | 309.5 | 315.4 | 319.7 | 311.5 | 319.3 | 286.6 | 295.3 |
| $\Delta AIC_{SRM} = (AIC_{SLRM} - AIC_{SRM})$ | 4.9 | -2.4 | 2.4 | -2.1 | 12 | 4.2 | 10 | 1.3 |

**Note:** \* $p < 0.1$ ; \*\* $p < 0.05$ ; \*\*\* $p < 0.01$ .

Controlling for spatial dependency in the residuals does not improve the results to the same extent. We find that there is evidence to favour the SEM specification only for 7 April. Nevertheless, the SLM models still offer a great improvement relative to the linear regression model. These results indicate that it is more important to account for spatial spill-over effects in mobility reduction than to control for spatial dependencies due to unobserved independent variables.

### 3.4 Spatial complexity II: The relevance of spatial heterogeneity

The SLM and SEM specifications remain global models that cannot fully capture variations in the associations of mobility reduction with SES, accessibility, activity commitments and public health at sub-national levels. The fact that the values for Δ*AIC*_*GWR*_ and 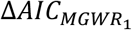(Table 4) are markedly greater than for Δ*AIC*_*SRM*_ indicates that considering spatial heterogeneity is more important than accounting for spatial dependency in analyses of the extent of mobility reduction during England’s lockdown. Together the values of the adjusted *R*^2^, Δ*AIC* and Δ*AIC*_*SRM*_ indicates that GWR and MGWR specifications clearly outperform the OLS specification for all four days. The values for Δ*AIC*, and Δ*AIC*_*SRM*_ are clearly >10, which means there is “essentially no support” (Burnham and Anderson, 2004, p. 71) for the SLRM specification.

The Δ*AIC* and Δ*AIC*_*c*_ values indicate that the MGWR specification is also superior to its GWR counterpart on 7 April. The case is less clear-cut for the other three dates although the Δ*AIC* and Δ*AIC*_*SRM*_ marginally favour the MGRW models, which provide valuable information on the scale at which different processes operate. Determining that scale as part of the modelling rather than deciding it a priori is useful since Table 4 shows that for three out of six regressors – non-English speakers, social grade C1 and poor health – the estimated bandwidth equals 190 and is thus estimated at the England-wide level on all days considered. It is the share of households in the top income quintile and resident population density – the two most important regressors– for which effects are more spatially restricted. The models fit the data best in East England, mainly Norfolk and surroundings, followed by the South East; adjusted *R*^2^ values are substantially lower in North England, typically more than 20 percent points than in the Norfolk and surrounding areas (Figure 4).

**Figure 4.**
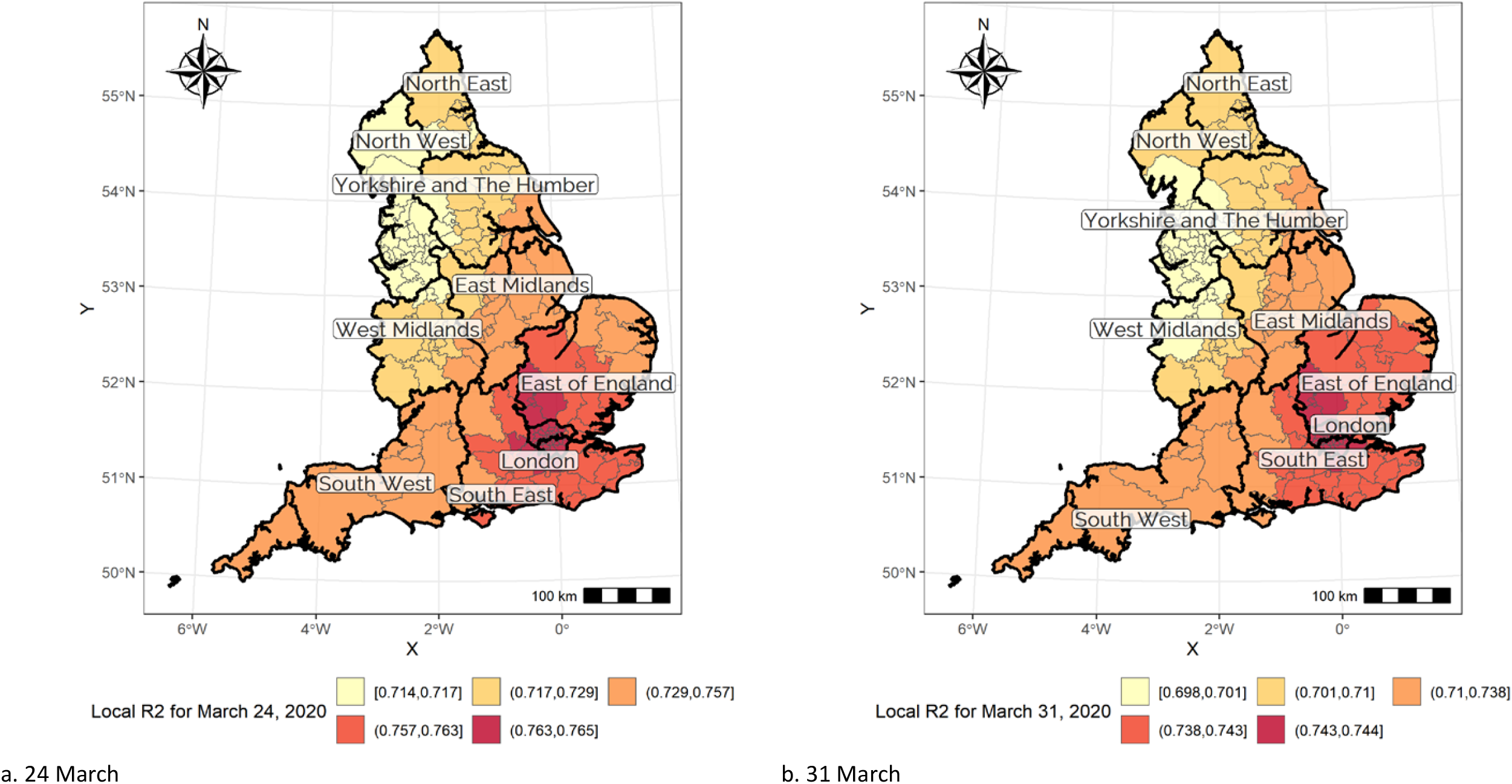
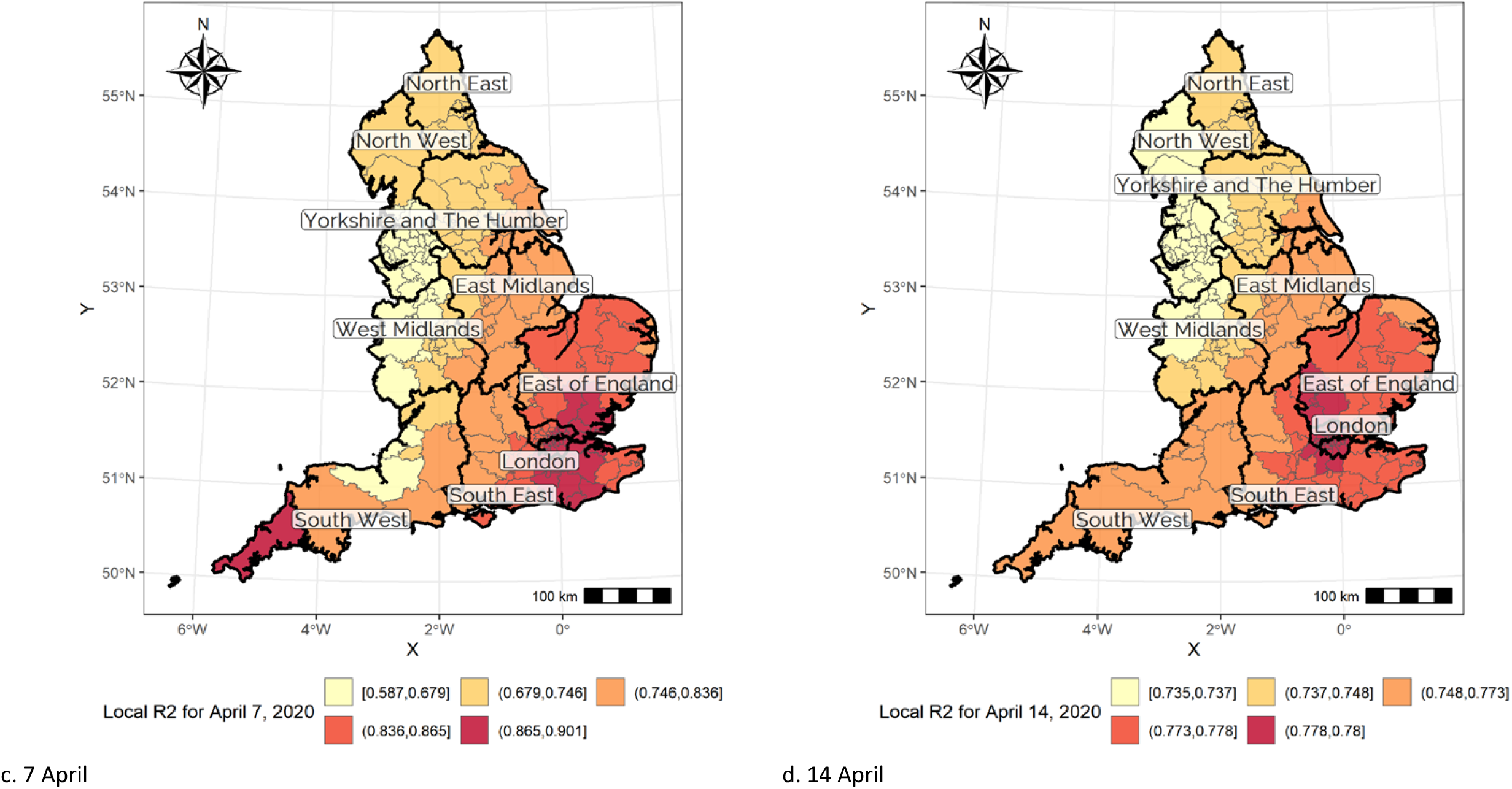
Local *R*^**2**^ in the MGWR model, by date.

The mean effects for the six regressors (Table 4) are stable over the four dates considered and broadly comparable to those for the SLRM specification. The model for 7 April tends to exhibit the greatest differences from other days when attention is directed towards the mean values. In addition, variation around the estimated mean value is markedly larger for the households in the top income quintile, residential density and the intercept. This indicates that spatial variation in the strength of the association with mobility reduction is most pronounced for the two strongest correlates plus the intercept. In relative terms, the variation around the mean is largest on three out of four days for residential density, but this is due to CCG areas with unusually high coefficients in the Lincoln-Hull region, followed by the North East. The right-skewed distribution of estimated coefficients can also be observed for households in the highest income quintile. Here the highest coefficients are observed for the area around Newcastle, followed by the rest of the North. In contrast, the lowest coefficients for the income and density variables can be found in South and East England. In those areas, the share of people in the lower middle classes (social grade C1) and with bad health are, relatively speaking, much more important to the explanation of mobility reduction. Greater London and its commuter belt also have high intercept values from 31 March onwards, suggesting that the base level of mobility reduction that is independent of any variable in the model has been rather high.

This discussion indicates substantial local variation in the correlates of mobility reduction in March and early April (Table 5). Consider, for instance, Newcastle upon Tyne in North East England for which the model explanatory power is comparatively low and spatial differences in mobility reduction are primarily a function of the share of households in the top quintile, residential density and the share of self-employed workers. This is different from London where there is a much more consistent base level reduction (i.e. intercept coefficient), and the explanatory power is distributed much more equally across all independent variables although bad health is relatively important and density and the self-employment less so. The implication of the local differences shown in Table 6 is that, whereas SES is important everywhere, the role of individual variables varies spatially.

## 4. Discussion

The reduction in everyday mobility across CCG areas in England in response to COVID-19 has been large, with a 70% reduction in the median radius of gyration, but broadly comparable with observations in other studies (Apple Inc, 2020; Google LLC, 2020; Jacobsen and Jacobsen, 2020; Pepe et al., 2020). These headline figures do not consider substantial variation at the individual or household level, which is where decisions about which trips to make or to forego are made. This is why the regression models presented in this article do not offer evidence of causal effects. Interpreting them in this manner would amount to ecological fallacy (Robinson, 1950). This does not, however, invalidate those models. After all, decisions about public health interventions as well as assessments of their effects and effectiveness take place at the level of populations and territories and not at the level of individuals or households.

The nature of the correlations of mobility reduction across England’s CCG areas with socio-economic status, accessibility, activity commitments and population health is in line with expectations. The strong influence of share of people in the top income quintile is not surprising, given that high-income workers tend to have greater discretion over when and where they work (Witteveen and Velthorst, 2020) and thus greater ability to work from home. In addition, people on high incomes tend to travel more than those on low incomes: individuals in the lowest household income quintile made 859 trips and covered 4,138 km in 2019 against 995 trips (+16%) and 9,236 km in the highest quintile (+223%) according to the UK National Travel Survey (Department for Transport, 2020b). The inclusion of the share of people in social grade C1 indicates that, once spatial differences in income are taken into account, CCG areas with more lower-middle workers experienced a lower total need to commute to/from work than territories with more working class individuals and households. Differences in ability to work from home are again relevant here, as is the observation that many key workers are employed in low-education, low-pay and precarious jobs associated with working class status in domains such as supermarkets, logistics and last-mile delivery, and construction.

The postive regression coefficients for the share of non-English speakers in Tables 2, 3 and 5 suggest that this variable may not be as much an indicator of SES as initially expected. As indicated above, the share of non-native English speakers is greatest in CCG areas where many people from non-Western heritage live. These individuals and households are comparatively likely to be excluded from car access and ownership (Lucas and Jones, 2009). They are thus strongly dependent on public transport, even when their concentration in higher-density locations (Badoe and Miller, 2000; Schimek, 1996) is taking into account. At the same time, it rapidly became clear over the spring of 2020 that ethnic minorities in the UK were at higher risk of COVID-19 infection and hospitalisation and mortality because of the virus (Proto and Quintana-Domeque, 2020). The coefficients in Tables 2, 3 and 5 may therefore also reflect a strong inclination to stay at home among groups from non-Western heritage because of perceived health risks.

Among the remaining variables, the postive coefficients for percent of the population in bad or very bad health and the negative coefficients for the share of population that is self-employed are as expected. The former most likely reflects a greater prevalence of government-encouraged shielding by clinically (extremely) vulnerable individuals and their households during the early stages of the epidemic. Yet, it may also reflect differences in local information campaigns, with health authorities in areas with large number of people with underlying health conditions perhaps placing greater emphasis on the need to shield and/or being more effective in reaching vulnerable individuals and households. It is also possible that greater and more effective community networks were in place or emerged in places with more clinically vulnerable households. Further research would be necessary to probe these conjectures. The result for share of self-employed workers seems to reflect autonomy over working hours and locations as well as discretion to engage in work-and business-related travel. Yet, a lack of choice also seems to be involved: for their business to survive and to make a living, many self-employed individuals may have to travel to suppliers and customers, whether they like it or not. Factors such as these may push up the (median) mobility at the CCG area level once differences in SES, accessibility and population health are taken into account.

The strong effect of population density is somewhat surprising but the positive correlation certainly not. Public transport use increases substantially with density (Badoe and Miller, 2000; Schimek, 1996) and this form of travel has been affected severely during the COVID-19 epidemic. Not only is it widely seen as a site where infection risk is particularly high, the capacity of service provision was dramatically reduced in the early stages of the epidemic and the Government’s official communication actively discouraged UK residents from using buses, trams, metros and trains. In addition, it is in high-density areas that the availability of local shops and delivery of groceries and other shopping bought online was most common before the epidemic (Brand et al., 2020). The need to travel for everyday needs is thus lower and can be satisfied locally more easily. Finally, the correlation of density with share of population of non-Western heritage may suggest a stronger inclination to stay at home due to perceived health risks in CCG areas with high population densities.

The above interpretations suggest that demographic compositions may be more important than geographic context at CCG area level. Nonetheless, neighbourhood factors may also hint at the effects of geographic context if there have indeed been differences in how CCGs have overseen the provision of spatially differentiated information to vulnerable individuals and their households. In addition, the relative prominence of compositional factors may reflect the scale at which mobility reductions have been analysed. The CCG level is an approriate choice in light of the healtcare sector’s response to the COVID-19 epidemic in England, but that does not mean that analysis conducted at another spatial level or using another classification of zones would have rendered the same results. The findings in this study remain subject to the modifiable areal unit problem (Fotheringham and Wong, 1991), and further work at different scales and with different spatial zones would enhance the understanding of how SES and other factors have shaped mobility reductions during the England-wide lockdown in early 2020.

The above analysis warrants two further conclusions. First, as the spatial regression models (Table 2) have suggested, mobility reduction in a given territory – even at the relatively aggregate level of the CCG area – is not independent from what happens in that territory’s neighbourhood. Figure 2 shows multiple, spatially extensive hot and cold, and the SLM specifications show that the spatially lagged dependent variables become statistically significant and meaningful regressors. Moreover, the inclusion of the latter reduces the coefficients of regressors measuring the attributes of the CCG area for which mobility reduction is analysed. This should come as a surprise: many people’s everyday mobility and activity spaces are not contained within the administrative territories on the basis of which healthcare services are provided. A comparison of Tables 3 suggests that SES, accessibility, activity commitments and population health in surrounding areas have shaped the extent of mobility reduction in a given CCG area. Had a spatial classification at a finer resolution been deployed, then the influence of neighbouring areas – and the need for spatially advanced econometrics – would have been even larger than in the present study.

Second, the relationships of mobility reduction with SES, accessibility, activity commitments and population health at the CCG level area vary significantly across England. As Table 3 demonstrates unambiguously, a spatially naïve linear regression model is simply not appropriate if the aim is to capture the spatial heterogeneity of the correlates of mobility reduction at the CCG area level. This is because on the one hand the relative importance of the regressors discussed above differs markedly across CCG areas, and on the other hand the underlying spatial processes play out over different spatial scales. The effects of the share of households in the top income quintile and resident population density – income and density – have the most pronounced geographies. Table 6 indicates that they have the strongest discriminatory effectsss in the post-industrial effects in North England, given that the coefficients for these variables are largest in Manchester, Leeds and Newcastle-on-Tyne while the overall goodness-of-fit (*R*^2^) for those cities is at most 87% of what it is in the City of London. These results are not altogether unexpected, given that spatial differences in the prevalence of high-income households (Figure 3) and resident population density are more pronounced across North England than across most of the North East.

**Table 3.**
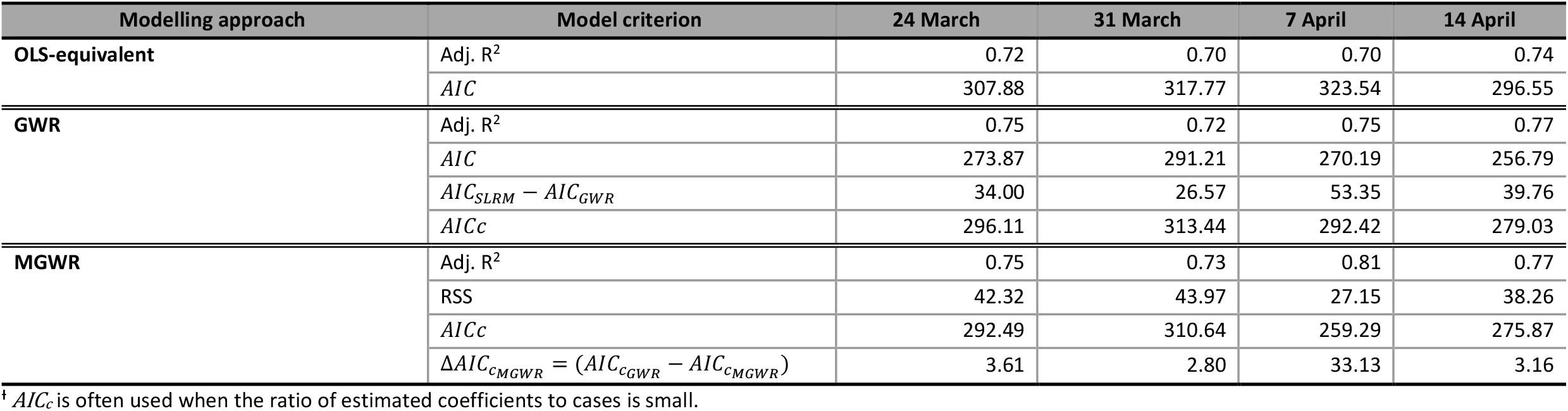
Assessment of spatial heterogeneity in regression modelling, by date.

| Modelling approach | Model criterion | 24 March | 31 March | 7 April | 14 April |
| --- | --- | --- | --- | --- | --- |
| **OLS-equivalent** | Adj. $R^2$ | 0.72 | 0.70 | 0.70 | 0.74 |
| | $AIC$ | 307.88 | 317.77 | 323.54 | 296.55 |
| **GWR** | Adj. $R^2$ | 0.75 | 0.72 | 0.75 | 0.77 |
| | $AIC$ | 273.87 | 291.21 | 270.19 | 256.79 |
| | $AIC_{SLRM} - AIC_{GWR}$ | 34.00 | 26.57 | 53.35 | 39.76 |
| | $AIC_c$ | 296.11 | 313.44 | 292.42 | 279.03 |
| **MGWR** | Adj. $R^2$ | 0.75 | 0.73 | 0.81 | 0.77 |
|  | RSS | 42.32 | 43.97 | 27.15 | 38.26 |
| | $AIC_c$ | 292.49 | 310.64 | 259.29 | 275.87 |
| | $\Delta AIC_{cMGWR} = (AIC_{cGWR} - AIC_{cMGWR})$ | 3.61 | 2.80 | 33.13 | 3.16 |

<sup>†</sup> $AIC_c$ is often used when the ratio of estimated coefficients to cases is small.

**Table 4A.**
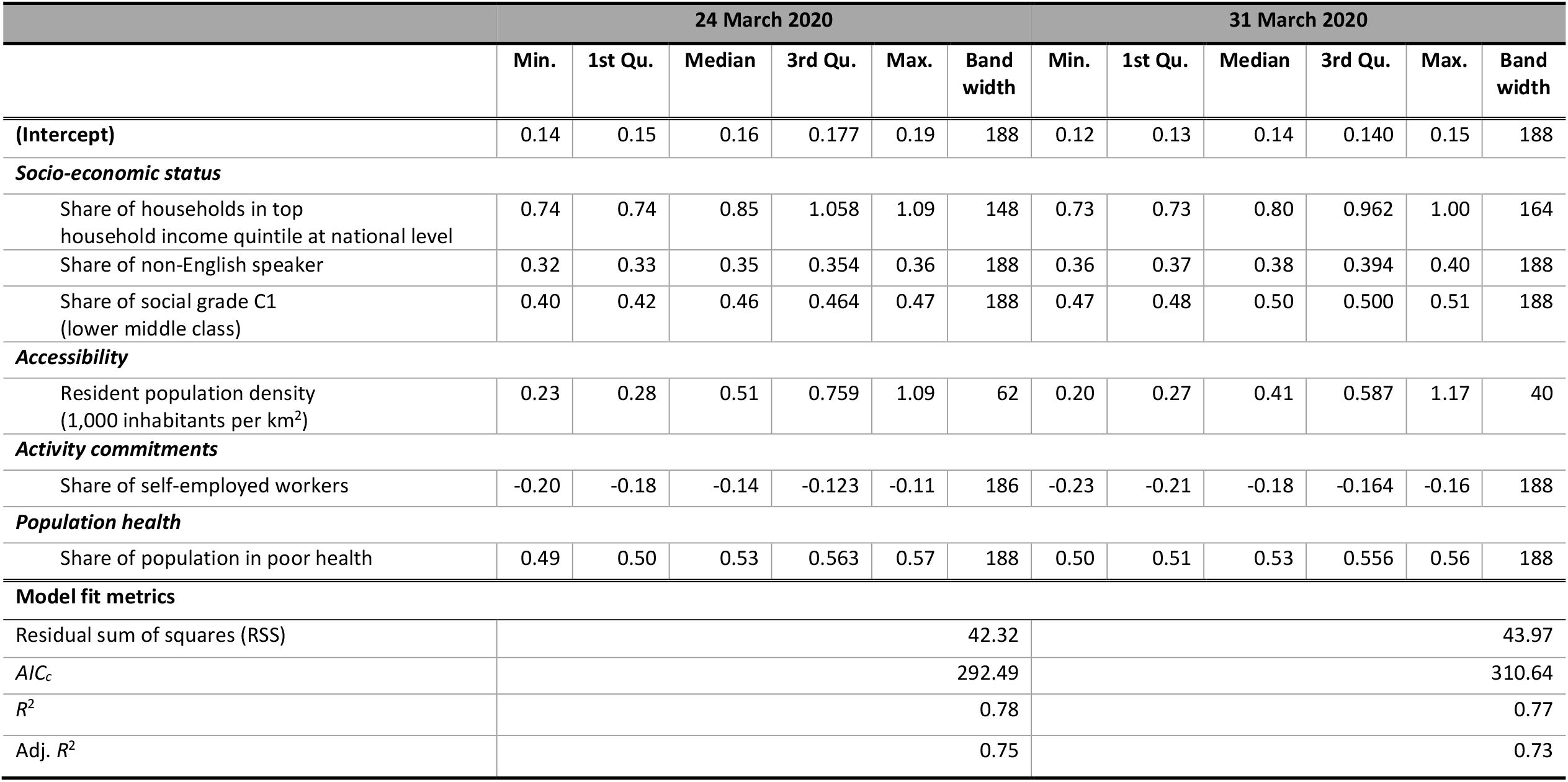
Results for MGWR modelling of mobility reduction, 24 and 31 March.

**Table 4B.**
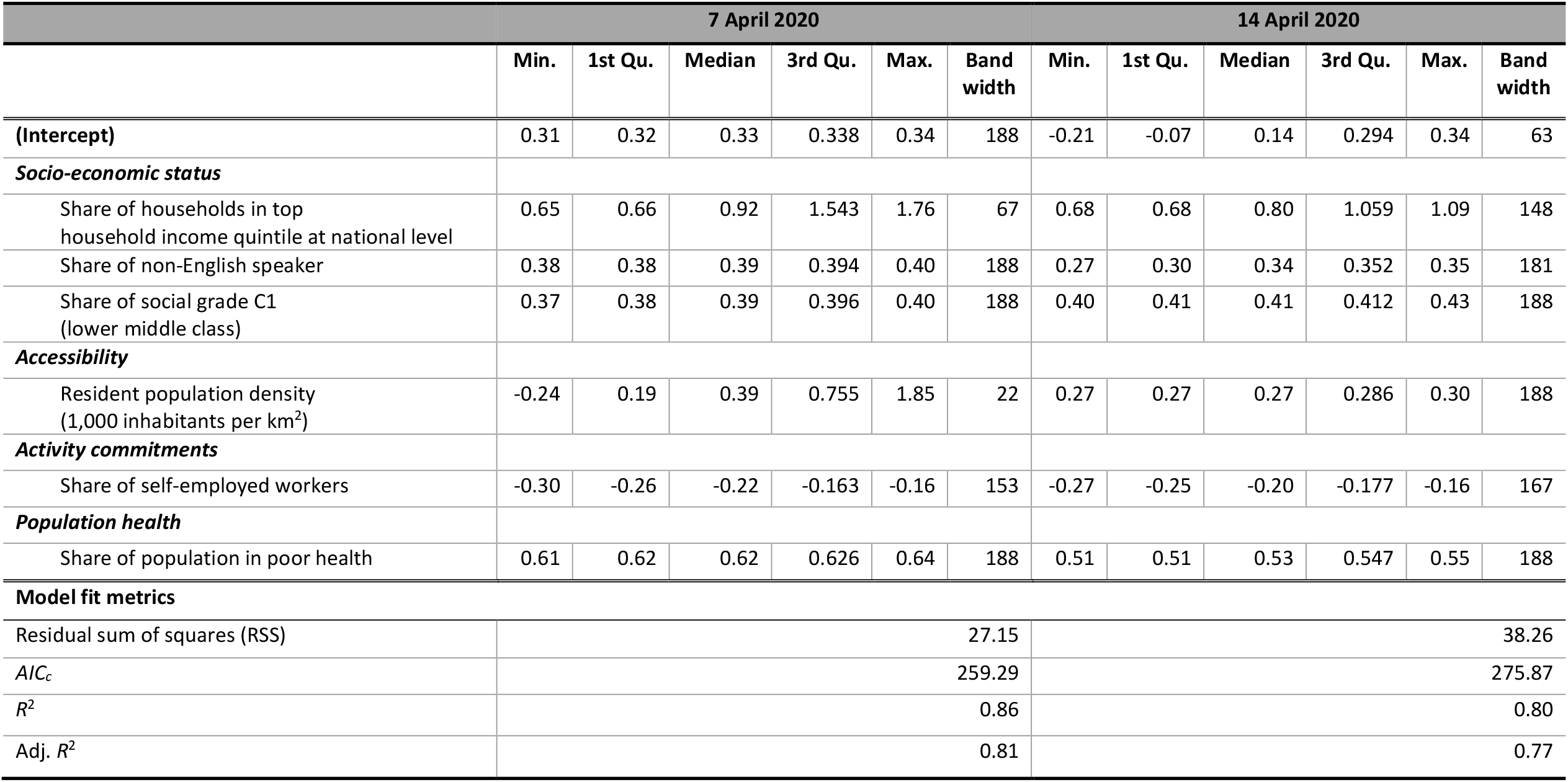
Results for MGWR modelling of mobility reduction, 7 and 14 April.

**Table 5.**
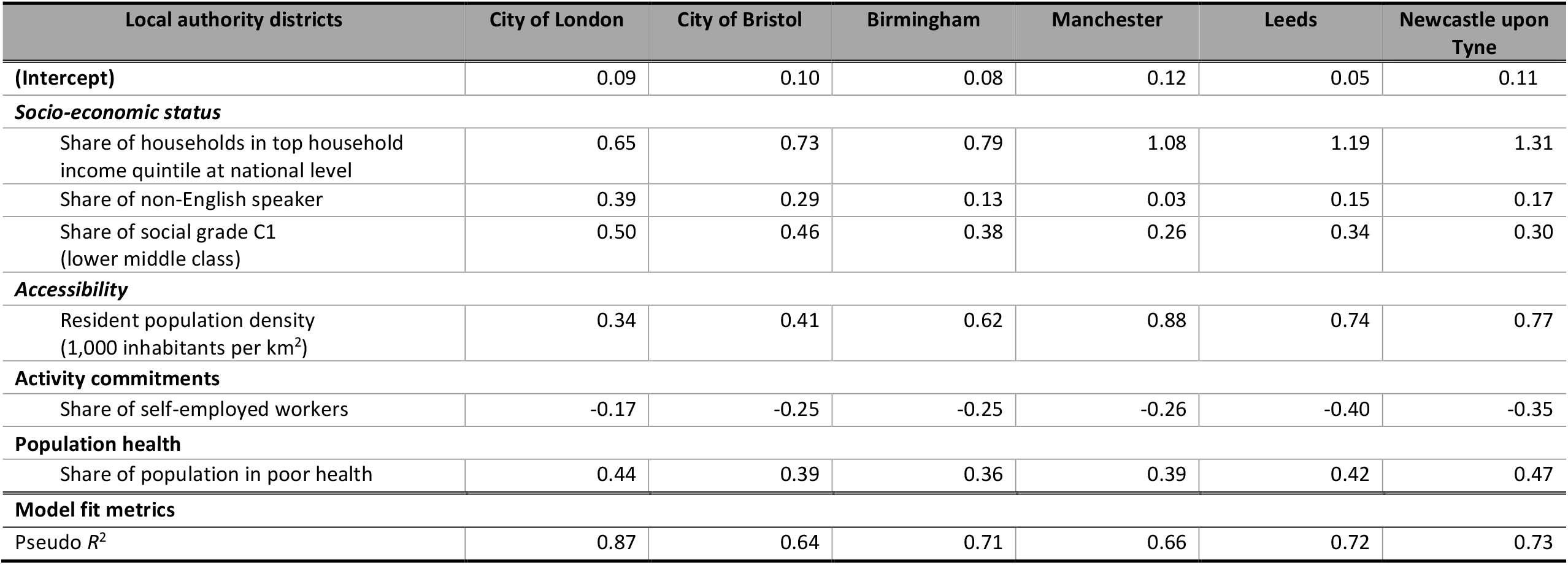
Locally specific coefficients in MGWR model, 7 April 2020.

| Local authority districts | City of London | City of Bristol | Birmingham | Manchester | Leeds | Newcastle upon Tyne |
| --- | --- | --- | --- | --- | --- | --- |
| **(Intercept)** | 0.09 | 0.10 | 0.08 | 0.12 | 0.05 | 0.11 |
| ***Socio-economic status*** |  |  |  |  |  |  |
| Share of households in top household income quintile at national level | 0.65 | 0.73 | 0.79 | 1.08 | 1.19 | 1.31 |
| Share of non-English speaker | 0.39 | 0.29 | 0.13 | 0.03 | 0.15 | 0.17 |
| Share of social grade C1 (lower middle class) | 0.50 | 0.46 | 0.38 | 0.26 | 0.34 | 0.30 |
| ***Accessibility*** |  |  |  |  |  |  |
| Resident population density (1,000 inhabitants per km<sup>2</sup>) | 0.34 | 0.41 | 0.62 | 0.88 | 0.74 | 0.77 |
| ***Activity commitments*** |  |  |  |  |  |  |
| Share of self-employed workers | -0.17 | -0.25 | -0.25 | -0.26 | -0.40 | -0.35 |
| ***Population health*** |  |  |  |  |  |  |
| Share of population in poor health | 0.44 | 0.39 | 0.36 | 0.39 | 0.42 | 0.47 |
| ***Model fit metrics*** |  |  |  |  |  |  |
| Pseudo $R^2$ | 0.87 | 0.64 | 0.71 | 0.66 | 0.72 | 0.73 |

## 5. Conclusion

This study has used mobile phone data to provide rigorous evidence for an link between socioeconomic status (SES) with mobility reduction during the Spring 2020 lockdown in England, while considering the potentially confounding effects of other factors and “recognising the fundamental spatiality of the current COVID-19 crisis” (Poom et al., 2020, p. 5). Two main conclusions can be drawn along with a future research direction.

First, SES has indeed been correlated with the extent of mobility reduction in the early stage of England’s COVID-19 epidemic. The headline figure of a 70% reduction in everyday mobility obscures marked spatial differences across local areas (Figure 1). All else equal, areas with more high-income households have seen the largest reductions in mobility, while those with more workers in lower middle-class occupations have seen greater reductions compared with those with more people in working-class occupations. The finding that areas with more non-English speakers have experienced greater reductions in mobility to some extent contravenes the suggestion that higher SES is associated with greater mobility reductions, but seems to reflect differences in the risk of infection, hospitalisation and death along lines of race/ethnicity. The greater reduction in mobility in territories with more non-English speakers appears to be an aggregate-level consequence of more shielding in response to the circulation of information about exposure risks, as it is in areas with more residents who are in bad or very bad health.

Second, the specific nature of the association of SES with mobility reduction varies markedly across England. There are both global (England-wide) and local factors that make a spatially naïve, conventional regression analysis inadequate and inappropriate. Territories as large as CCG areas are not independent as reductions in one area are influenced by those in surrounding areas due to everyday mobility patterns’ transgression of administrative boundaries. More importantly, the strength of correlations between mobility reduction on the one hand and SES, accessibility, activity commitments and population health differs markedly across the country. Additionally, the significance of SES is larger across Northern England than in London and the South East.

Our study leaves open the question why (economically) disenfranchised areas had a lower compliance with lockdowns. Moreover, one may conjecture that local areas in which many residents could not afford the relative luxury of staying at home and reducing their mobility as much as in more privileged areas are at a disadvantage (e.g., more infections, overwhelmed healthcare services) in later stages of the COVID-19. Verification of this conjecture is beyond the current study but the path-dependent spatial differentiation in ability to reduce everyday mobility is an important topic for future research.

## Data Availability

All source R code and data necessary for the replication of our results and figures are available at https://github.com/wondolee/COVID19.

## Availability of data and materials

All source R code and data necessary for the replication of our results and figures is available at https://github.com/wondolee/COVID19.

## Credit authorship contribution statement

Won Do Lee: Conceptualisation, Data Curation, Methodology, Formal analysis, Writing-Original draft preparation. Matthias Qian: Data Curation, Writing-Reviewing and Editing. Tim Schwanen: Conceptualisation, Writing-Original draft preparation, Supervision, Writing-Reviewing and Editing.

## Acknowledgement

The authors would like to thank CKDelta company as the project partner of “Oxford COVID-19 Impact Monitor”, to provide a wide array of anonymised and aggregated data. This research did not receive any specific grant from funding agencies in the public, commercial, or not-for-profit sectors.

## Footnotes

1 Two percent of the population has been sub-sampled from the users of a large British mobile phone provider, stratified by the 191 Clinical Commissioning Groups (the reconfiguration of CCGs have taken place in April 2020, a number of CCGs have reduced from 191 to 136 in April 2020) in England.

2 Using data from Qian et al. 2020, we find a 96% Pearson correlation between the proportion of users staying at home and the median radius of gyration for users across the United Kingdom, providing further evidence of the equivalence of measures of the range and amount of mobility.

3 The computation of home region of users exploits the night-time location when users are most likely to be at home. Home region detection followed three steps: a) filter observations from 10 pm to 6 am, b) finding the most common cell phone tower used at night-time, c) dropping users with fewer than 30 night-time observations per month. Each cell phone tower is assigned to its Clinical Commissioning Group area according to its location.

