## Supplementary Data for "The association between socioeconomic status and mobility reductions in the early stage of England’s COVID-19 epidemic"

Appendix A. Supplementary Data
Supplement Materials for

### The association between socioeconomic status and mobility reductions in the early stage of England's COVID-19 epidemic

This study estimated the mobility levels of all 191 clinical commissioning group (CCG) areas in England. As the measure of mobility, we use the daily median radius of gyration[1]. To examine how socioeconomic status was associated with the extent of mobility reduction during the Spring 2020 lockdown in England, we considered potentially confounding effects and spatial dependency and heterogeneity to construct the model specification. We then applied spatial regression models that account for global and local spatial autocorrelation. Finally, we compared measures of model fit among the developed models by considering the adjusted $R^2$, residual sum of squares (RSS), $\Delta AIC$ and $\Delta AICc.$ We found that the GWR and MGWR specifications outperform the OLS specification on four consecutive Tuesdays starting on 24 March.

A potential concern for our empirical results is the choice of our reference period. Our main results compare the mobility on Tuesdays after the lockdown with Tuesday 3 March 2020. Instead of considering reference days, we can also consider reference weeks. In this appendix, we show the results of two robustness checks involving weekly references. First, we can compare mobility on Tuesdays following the national lockdown with the weekly reference (3 March 2020 ± 3 days). Second, we can compare the weekly reduction in mobility following the national lockdown with the conformable reference week.

Tables 1 to 6 reproduce the results from our main manuscript, where we use daily reference periods. Tables 7 to 12 present the comparison of post-lockdown mobility reductions on Tuesdays to the weekly reference. Tables 13 to 18 show the comparison of lost-lockdown weekly reductions in mobility to the weekly reference. Overall, the differences are small. The selected variables with daily reference periods remain highly significant when weekly reference periods are used. The measures of fit do not consistently favour daily over weekly reference periods or vice versa. We conclude that our findings are robust to the choice of the reference period.

[1] González, M.C., Hidalgo, C.A., Barabási, A.-L., 2008. Understanding individual human mobility patterns. *Nature* 453, 779–782. https://doi.org/10.1038/nature06958

#### Measuring mobility reductions of each day, compared to the baseline day.

- A comparison between the median radius of gyration across England on selected post-lockdown Tuesdays and the baseline (3 March 2020) mobility levels.
- Table 1 presents the summary of model fit metrics among the developed models using the measured mobility levels compared to the baseline day.
- Table 2 shows the estimated coefficients of step-wise linear regression models (ordinary least squares, hereafter OLS) by date.
- Table 3 to Table 6 show the range of estimated coefficients and model fit metrics for the GWR and MGWR models by date.

**Table 1. Assessment of spatial heterogeneity in regression modelling, the dependent variable was estimated by comparing the mobility level on each of the selected post-lockdown Tuesdays with baseline (3 March 2020) estimates.**

| Modelling approach | Model criterion | 24 March | 31 March | 7 April | 14 April |
|---|---|---|---|---|---|
| OLS | Adj. $R^2$ | 0.72 | 0.70 | 0.70 | 0.74 |
| | $AIC$ | 307.88 | 317.77 | 323.54 | 296.55 |
| GWR | Adj. $R^2$ | 0.75 | 0.72 | 0.75 | 0.77 |
| | $AIC$ | 273.87 | 291.21 | 270.19 | 256.79 |
| | $AIC_{SLRM} - AIC_{GWR}$ | 34.00 | 26.57 | 53.35 | 39.76 |
| | $AICc$ | 296.11 | 313.44 | 292.42 | 279.03 |
| MGWR | Adj. $R^2$ | 0.75 | 0.73 | 0.81 | 0.77 |
| | RSS | 42.32 | 43.97 | 27.15 | 38.26 |
| | $AICc$ | 292.49 | 310.64 | 259.29 | 275.87 |
| | $\Delta AIC_{c_{MGWR}} = (AIC_{c_{GWR}} - AIC_{c_{MGWR}})$ | 3.61 | 2.80 | 33.13 | 3.16 |

Note: GWR and MGWR both uses the bi-square Kernel function and calibrates the bandwidth on the basis of *k*=4 nearest neighbours to generate the local weightings.

**Table 2. Step-wise linear regression model for mobility reduction, by date.**

| Mobility measured compare to the baseline | 24 March 2020 | | 31 March 2020 | | 7 April 2020 | | 14 April 2020 | |
|---|---|---|---|---|---|---|---|---|
| | Coeff (SD) | LMG[2] | Coeff (SD) | LMG | Coeff (SD) | LMG | Coeff (SD) | LMG |
| **(Intercept)** | 0.00 (0.04) | - | 0.00 (0.04) | - | 0.00 (0.04) | - | 0.000 (0.04) | - |
| ***Socio-economic status*** | | | | | | | | |
| Share of households in top household income quintile at national level | 0.80*** (0.07) | 0.34 | 0.81*** (0.07) | 0.33 | 0.78*** (0.07) | 0.34 | 0.79*** (0.07) | 0.34 |
| Share of non-English speaker | 0.36*** (0.07) | 0.17 | 0.40*** (0.07) | 0.18 | 0.33*** (0.07) | 0.17 | 0.31*** (0.07) | 0.15 |
| Share of social grade C1 (lower middle class) | 0.51*** (0.06) | 0.08 | 0.54*** (0.06) | 0.09 | 0.50*** (0.06) | 0.08 | 0.53*** (0.06) | 0.09 |
| ***Accessibility*** | | | | | | | | |
| Resident population density (1,000 inhabitants per $km^2$) | 0.37*** (0.08) | 0.29 | 0.35*** (0.08) | 0.27 | 0.40*** (0.08) | 0.29 | 0.42*** (0.08) | 0.28 |
| ***Activity commitments*** | | | | | | | | |
| Share of self-employed workers | -0.21*** (0.06) | 0.05 | -0.25*** (0.06) | 0.05 | -0.27*** (0.07) | 0.05 | -0.22*** (0.06) | 0.06 |
| ***Population health*** | | | | | | | | |
| Share of population in bad health | 0.46*** (0.07) | 0.07 | 0.49*** (0.08) | 0.08 | 0.41*** (0.08) | 0.07 | 0.43*** (0.07) | 0.07 |
| **Model fit metrics** | | | | | | | | |
| $R^2$ | 0.73 | | 0.71 | | 0.71 | | 0.74 | |
| Adjusted $R^2$ | 0.72 | | 0.71 | | 0.70 | | 0.74 | |
| Residual Std. Error (df = 184) | 0.53 | | 0.54 | | 0.55 | | 0.51 | |
| F Statistic (df = 6; 184) | 82.36*** | | 76.66*** | | 73.5*** | | 89.27*** | |

[2] Relative importance metric, the contribution of each variable of $R^2$ (Lindeman et al., 1980).

**Note: *$p$<0.1; **$p$<0.05; ***$p$<0.01.**

**Table 3. Results for GWR modelling of mobility reduction, 24 and 31 March.**

| Mobility measured compare to the baseline | 24 March 2020 | | | | | | 31 March 2020 | | | | | |
|---|---|---|---|---|---|---|---|---|---|---|---|---|
| | Min. | 1st Qu. | Median | 3rd Qu. | Max. | Band width | Min. | 1st Qu. | Median | 3rd Qu. | Max. | Band width |
| **(Intercept)** | -0.10 | 0.03 | 0.04 | 0.119 | 0.19 | 155 | -0.10 | 0.02 | 0.05 | 0.108 | 0.16 | 155 |
| ***Socio-economic status*** | | | | | | | | | | | | |
| Share of households in top household income quintile at national level | 0.65 | 0.67 | 0.797 | 1.06 | 1.25 | 155 | 0.63 | 0.67 | 0.78 | 1.041 | 1.23 | 155 |
| Share of non-English speaker | 0.22 | 0.31 | 0.343 | 0.36 | 0.46 | 155 | 0.29 | 0.36 | 0.37 | 0.389 | 0.44 | 155 |
| Share of social grade C1 (lower middle class) | 0.36 | 0.43 | 0.511 | 0.52 | 0.62 | 155 | 0.46 | 0.50 | 0.54 | 0.551 | 0.64 | 155 |
| ***Accessibility*** | | | | | | | | | | | | |
| Resident population density (1,000 inhabitants per $km^2$) | 0.32 | 0.37 | 0.397 | 0.63 | 0.73 | 155 | 0.34 | 0.36 | 0.39 | 0.557 | 0.62 | 155 |
| ***Activity commitments*** | | | | | | | | | | | | |
| Share of self-employed workers | -0.48 | -0.27 | -0.168 | -0.08 | -0.05 | 155 | -0.51 | -0.32 | -0.21 | -0.128 | -0.07 | 155 |
| ***Population health*** | | | | | | | | | | | | |
| Share of population in poor health | 0.41 | 0.44 | 0.458 | 0.48 | 0.57 | 155 | 0.44 | 0.45 | 0.47 | 0.490 | 0.58 | 155 |
| **Model fit metrics** | | | | | | | | | | | | |
| **Residual sum of squares (RSS)** | | | | | | 43.03 | | | | | | 47.12 |
| ***AIC*** | | | | | | 273.87 | | | | | | 291.21 |
| $AIC_c$ | | | | | | 296.11 | | | | | | 313.44 |
| $R^2$ | | | | | | 0.77 | | | | | | 0.75 |
| **Adj.** $R^2$ | | | | | | 0.75 | | | | | | 0.72 |

**Table 4. Results for GWR modelling of mobility reduction, 7 and 14 April.**

| Mobility measured compare to the baseline | 7 April 2020 | | | | | | 14 April 2020 | | | | | |
|---|---|---|---|---|---|---|---|---|---|---|---|---|
| | Min. | 1st Qu. | Median | 3rd Qu. | Max. | Band width | Min. | 1st Qu. | Median | 3rd Qu. | Max. | Band width |
| **(Intercept)** | -0.11 | 0.07 | 0.09 | 0.115 | 0.17 | 155 | -0.07 | 0.04 | 0.06 | 0.074 | 0.11 | 155 |
| ***Socio-economic status*** | | | | | | | | | | | | |
| Share of households in top household income quintile at national level | 0.62 | 0.65 | 0.73 | 1.074 | 1.33 | 155 | 0.61 | 0.65 | 0.79 | 1.060 | 1.20 | 155 |
| Share of non-English speaker | -0.01 | 0.10 | 0.35 | 0.391 | 0.44 | 155 | 0.04 | 0.10 | 0.31 | 0.379 | 0.39 | 155 |
| Share of social grade C1 (lower middle class) | 0.24 | 0.31 | 0.48 | 0.497 | 0.58 | 155 | 0.35 | 0.40 | 0.50 | 0.505 | 0.55 | 155 |
| ***Accessibility*** | | | | | | | | | | | | |
| Resident population density (1,000 inhabitants per $km^2$) | 0.31 | 0.33 | 0.41 | 0.774 | 0.89 | 155 | 0.34 | 0.36 | 0.40 | 0.694 | 0.76 | 155 |
| ***Activity commitments*** | | | | | | | | | | | | |
| Share of self-employed workers | -0.65 | -0.27 | -0.23 | -0.175 | -0.13 | 155 | -0.46 | -0.29 | -0.19 | -0.122 | -0.05 | 155 |
| ***Population health*** | | | | | | | | | | | | |
| Share of population in poor health | 0.33 | 0.40 | 0.43 | 0.448 | 0.49 | 155 | 0.37 | 0.40 | 0.42 | 0.435 | 0.55 | 155 |
| **Model fit metrics** | | | | | | | | | | | | |
| **Residual sum of squares (RSS)** | 42.21 | | | | | | 39.35 | | | | | |
| ***AIC*** | 270.19 | | | | | | 256.79 | | | | | |
| $AIC_c$ | 292.42 | | | | | | 279.03 | | | | | |
| $R^2$ | 0.78 | | | | | | 0.79 | | | | | |
| **Adj.** $R^2$ | 0.75 | | | | | | 0.77 | | | | | |

**Table 5. Results for MGWR modelling of mobility reduction, 24 and 31 March.**

| Mobility measured compare to the baseline | 24 March 2020 | | | | | | 31 March 2020 | | | | | |
|---|---|---|---|---|---|---|---|---|---|---|---|---|
| | Min. | 1st Qu. | Median | 3rd Qu. | Max. | Band width | Min. | 1st Qu. | Median | 3rd Qu. | Max. | Band width |
| **(Intercept)** | 0.14 | 0.15 | 0.16 | 0.177 | 0.19 | 188 | 0.12 | 0.13 | 0.14 | 0.140 | 0.15 | 188 |
| ***Socio-economic status*** | | | | | | | | | | | | |
| Share of households in top household income quintile at national level | 0.74 | 0.74 | 0.85 | 1.058 | 1.09 | 148 | 0.73 | 0.73 | 0.80 | 0.962 | 1.00 | 164 |
| Share of non-English speaker | 0.32 | 0.33 | 0.35 | 0.354 | 0.36 | 188 | 0.36 | 0.37 | 0.38 | 0.394 | 0.40 | 188 |
| Share of social grade C1 (lower middle class) | 0.40 | 0.42 | 0.46 | 0.464 | 0.47 | 188 | 0.47 | 0.48 | 0.50 | 0.500 | 0.51 | 188 |
| ***Accessibility*** | | | | | | | | | | | | |
| Resident population density (1,000 inhabitants per $km^2$) | 0.23 | 0.28 | 0.51 | 0.759 | 1.09 | 62 | 0.20 | 0.27 | 0.41 | 0.587 | 1.17 | 40 |
| ***Activity commitments*** | | | | | | | | | | | | |
| Share of self-employed workers | -0.20 | -0.18 | -0.14 | -0.123 | -0.11 | 186 | -0.23 | -0.21 | -0.18 | -0.164 | -0.16 | 188 |
| ***Population health*** | | | | | | | | | | | | |
| Share of population in poor health | 0.49 | 0.50 | 0.53 | 0.563 | 0.57 | 188 | 0.50 | 0.51 | 0.53 | 0.556 | 0.56 | 188 |
| **Model fit metrics** | | | | | | | | | | | | |
| **Residual sum of squares (RSS)** | 42.32 | | | | | | 43.97 | | | | | |
| $AIC_c$ | 292.49 | | | | | | 310.64 | | | | | |
| $R^2$ | 0.78 | | | | | | 0.77 | | | | | |
| **Adj.** $R^2$ | 0.75 | | | | | | 0.73 | | | | | |

**Table 6. Results for MGWR modelling of mobility reduction, 7 and 14 April.**

| Mobility measured compare to the baseline | 7 April 2020 | | | | | | 14 April 2020 | | | | | |
|---|---|---|---|---|---|---|---|---|---|---|---|---|
| | Min. | 1st Qu. | Median | 3rd Qu. | Max. | Band width | Min. | 1st Qu. | Median | 3rd Qu. | Max. | Band width |
| **(Intercept)** | 0.31 | 0.32 | 0.33 | 0.338 | 0.34 | 188 | -0.21 | -0.07 | 0.14 | 0.294 | 0.34 | 63 |
| ***Socio-economic status*** | | | | | | | | | | | | |
| Share of households in top household income quintile at national level | 0.65 | 0.66 | 0.92 | 1.543 | 1.76 | 67 | 0.68 | 0.68 | 0.80 | 1.059 | 1.09 | 148 |
| Share of non-English speaker | 0.38 | 0.38 | 0.39 | 0.394 | 0.40 | 188 | 0.27 | 0.30 | 0.34 | 0.352 | 0.35 | 181 |
| Share of social grade C1 (lower middle class) | 0.37 | 0.38 | 0.39 | 0.396 | 0.40 | 188 | 0.40 | 0.41 | 0.41 | 0.412 | 0.43 | 188 |
| ***Accessibility*** | | | | | | | | | | | | |
| Resident population density (1,000 inhabitants per km$^2$) | -0.24 | 0.19 | 0.39 | 0.755 | 1.85 | 22 | 0.27 | 0.27 | 0.27 | 0.286 | 0.30 | 188 |
| ***Activity commitments*** | | | | | | | | | | | | |
| Share of self-employed workers | -0.30 | -0.26 | -0.22 | -0.163 | -0.16 | 153 | -0.27 | -0.25 | -0.20 | -0.177 | -0.16 | 167 |
| ***Population health*** | | | | | | | | | | | | |
| Share of population in poor health | 0.61 | 0.62 | 0.62 | 0.626 | 0.64 | 188 | 0.51 | 0.51 | 0.53 | 0.547 | 0.55 | 188 |
| **Model fit metrics** | | | | | | | | | | | | |
| **Residual sum of squares (RSS)** | 27.15 | | | | | | 38.26 | | | | | |
| ***$AIC_c$*** | 259.29 | | | | | | 275.87 | | | | | |
| ***$R^2$*** | 0.86 | | | | | | 0.80 | | | | | |
| **Adj. $R^2$** | 0.81 | | | | | | 0.77 | | | | | |

#### Measuring mobility reductions of each day, compared to the baseline week.

- A comparison between the median radius of gyration across England for selected post-lockdown Tuesdays and the baseline week (3 March 2020 ± 3 days) mobility levels.
- Table 7 presents the summary of model fit metrics among the developed models using the measured mobility levels compared to the baseline week.
- Table 8 shows the estimated coefficients of OLS models by date.
- Table 9 to Table 12 show the range of estimated coefficients and model fit metrics for the GWR and MGWR models by date.

**Table 7. Assessment of spatial heterogeneity in regression modelling, the dependent variable was estimated by comparing the mobility level on each of the selected post-lockdown Tuesdays with baseline week (3 March 2020 ± 3 days) estimates.**

| Modelling approach | Model criterion | 24 March | 31 March | 7 April | 14 April |
|---|---|---|---|---|---|
| **OLS** | Adj. $R^2$ | 0.66 | 0.67 | 0.66 | 0.69 |
| | $AIC$ | 343.95 | 336.87 | 343.34 | 328.39 |
| **GWR** | Adj. $R^2$ | 0.79 | 0.82 | 0.82 | 0.83 |
| | $AIC$ | 221.38 | 200.58 | 200.46 | 187.57 |
| | $AIC_{SLRM} - AIC_{GWR}$ | 122.57 | 136.28 | 142.88 | 140.82 |
| | $AICc$ | 280.93 | 260.13 | 260.01 | 247.12 |
| **MGWR** | Adj. $R^2$ | 0.79 | 0.81 | 0.83 | 0.84 |
| | RSS | 32.62 | 26.63 | 23.80 | 22.06 |
| | $AICc$ | 279.82 | 269.73 | 255.94 | 247.78 |
| | $\Delta AIC_{c_{MGWR}} = (AIC_{c_{GWR}} - AIC_{c_{MGWR}})$ | 1.11 | -9.60 | 4.07 | -0.66 |

Note: GWR and MGWR both uses the bi-square Kernel function and calibrates the bandwidth on the basis of *k*=4 nearest neighbours to generate the local weightings.

**Table 8. Step-wise linear regression model for mobility reduction, by date.**

| Mobility measured compare to the baseline week | 24 March 2020 | | 31 March 2020 | | 7 April 2020 | | 14 April 2020 | |
|---|---|---|---|---|---|---|---|---|
| | Coeff (SD) | LMG | Coeff (SD) | LMG | Coeff (SD) | LMG | Coeff (SD) | LMG |
| **(Intercept)** | 0.00 (0.04) | - | 0.00 (0.04) | - | 0.00 (0.04) | - | 0.000 (0.04) | - |
| ***Socio-economic status*** | | | | | | | | |
| Share of households in top household income quintile at national level | 0.61*** (0.08) | 0.17 | 0.61*** (0.08) | 0.16 | 0.61*** (0.08) | 0.18 | 0.61*** (0.08) | 0.17 |
| Share of non-English speaker | 0.44*** (0.08) | 0.27 | 0.46*** (0.08) | 0.27 | 0.44*** (0.08) | 0.26 | 0.44*** (0.07) | 0.26 |
| Share of social grade C1 (lower middle class) | 0.34*** (0.07) | 0.08 | 0.35*** (0.07) | 0.08 | 0.34*** (0.07) | 0.08 | 0.35*** (0.06) | 0.08 |
| ***Accessibility*** | | | | | | | | |
| Resident population density (1,000 inhabitants per km$^2$) | 0.39*** (0.09) | 0.33 | 0.38*** (0.09) | 0.33 | 0.40*** (0.09) | 0.33 | 0.42*** (0.08) | 0.34 |
| ***Activity commitments*** | | | | | | | | |
| Share of self-employed workers | -0.58*** (0.07) | 0.11 | -0.60*** (0.07) | 0.12 | -0.60*** (0.07) | 0.12 | -0.60*** (0.07) | 0.11 |
| ***Population health*** | | | | | | | | |
| Share of population in bad health | 0.29*** (0.08) | 0.04 | 0.31*** (0.08) | 0.04 | 0.27*** (0.08) | 0.04 | 0.28*** (0.08S) | 0.04 |
| **Model fit metrics** | | | | | | | | |
| $R^2$ | | 0.73 | | 0.71 | | 0.71 | | 0.74 |
| Adjusted $R^2$ | | 0.72 | | 0.71 | | 0.70 | | 0.74 |
| Residual Std. Error (df = 184) | | 0.53 | | 0.54 | | 0.55 | | 0.51 |
| F Statistic (df = 6; 184) | | 82.36*** | | 76.66*** | | 73.46*** | | 89.27*** |

**Note: *$p$<0.1; **$p$<0.05; ***$p$<0.01.**

**Table 9. Results for GWR modelling of mobility reduction, 24 and 31 March.**

| Mobility measured compare to the baseline week | 24 March 2020 | | | | | | 31 March 2020 | | | | | |
|---|---|---|---|---|---|---|---|---|---|---|---|---|
| | Min. | 1st Qu. | Median | 3rd Qu. | Max. | Band width | Min. | 1st Qu. | Median | 3rd Qu. | Max. | Band width |
| **(Intercept)** | -0.25 | 0.12 | 0.22 | 0.434 | 0.59 | 73 | -0.29 | 0.12 | 0.21 | 0.392 | 0.58 | 73 |
| ***Socio-economic status*** | | | | | | | | | | | | |
| Share of households in top household income quintile at national level | 0.29 | 0.41 | 0.683 | 1.12 | 1.58 | 73 | 0.18 | 0.37 | 0.70 | 1.152 | 1.55 | 73 |
| Share of non-English speaker | 0.07 | 0.30 | 0.360 | 0.70 | 0.97 | 73 | 0.02 | 0.30 | 0.40 | 0.788 | 0.99 | 73 |
| Share of social grade C1 (lower middle class) | 0.06 | 0.20 | 0.311 | 0.43 | 0.62 | 73 | 0.08 | 0.21 | 0.37 | 0.478 | 0.69 | 73 |
| ***Accessibility*** | | | | | | | | | | | | |
| Resident population density (1,000 inhabitants per km$^2$) | 0.14 | 0.24 | 0.678 | 1.21 | 1.32 | 73 | 0.02 | 0.23 | 0.62 | 1.081 | 1.46 | 73 |
| ***Activity commitments*** | | | | | | | | | | | | |
| Share of self-employed workers | -1.26 | -0.74 | -0.639 | -0.18 | -0.11 | 73 | -1.27 | -0.79 | -0.68 | -0.166 | -0.09 | 73 |
| ***Population health*** | | | | | | | | | | | | |
| Share of population in poor health | -0.07 | 0.19 | 0.241 | 0.35 | 0.76 | 73 | -0.08 | 0.16 | 0.25 | 0.401 | 0.98 | 73 |
| **Model fit metrics** | | | | | | | | | | | | |
| **Residual sum of squares (RSS)** | 29.29 | | | | | | 26.27 | | | | | |
| ***AIC*** | 221.38 | | | | | | 200.58 | | | | | |
| ***AIC$_c$*** | 280.93 | | | | | | 260.13 | | | | | |
| $R^2$ | 0.85 | | | | | | 0.86 | | | | | |
| **Adj.** $R^2$ | 0.79 | | | | | | 0.82 | | | | | |

**Table 10. Results for GWR modelling of mobility reduction, 7 and 14 April.**

| Mobility measured compare to the baseline week | 7 April 2020 | | | | | | 14 April 2020 | | | | | |
|---|---|---|---|---|---|---|---|---|---|---|---|---|
| | Min. | 1st Qu. | Median | 3rd Qu. | Max. | Band width | Min. | 1st Qu. | Median | 3rd Qu. | Max. | Band width |
| **(Intercept)** | -0.25 | 0.14 | 0.25 | 0.394 | 0.50 | 73 | -0.25 | 0.14 | 0.24 | 0.340 | 0.51 | 73 |
| ***Socio-economic status*** | | | | | | | | | | | | |
| Share of households in top household income quintile at national level | 0.21 | 0.40 | 0.72 | 1.065 | 1.90 | 73 | 0.27 | 0.46 | 0.70 | 1.116 | 1.44 | 73 |
| Share of non-English speaker | 0.09 | 0.31 | 0.37 | 0.534 | 1.10 | 73 | -0.01 | 0.32 | 0.37 | 0.590 | 0.86 | 73 |
| Share of social grade C1 (lower middle class) | 0.07 | 0.20 | 0.30 | 0.392 | 0.64 | 73 | 0.03 | 0.18 | 0.26 | 0.421 | 0.69 | 73 |
| ***Accessibility*** | | | | | | | | | | | | |
| Resident population density (1,000 inhabitants per $km^2$) | 0.13 | 0.20 | 0.55 | 1.206 | 1.39 | 73 | 0.11 | 0.19 | 0.65 | 1.103 | 1.33 | 73 |
| ***Activity commitments*** | | | | | | | | | | | | |
| Share of self-employed workers | -1.39 | -0.75 | -0.59 | -0.183 | -0.10 | 73 | -1.27 | -0.78 | -0.66 | -0.208 | -0.11 | 73 |
| ***Population health*** | | | | | | | | | | | | |
| Share of population in poor health | -0.07 | 0.19 | 0.27 | 0.365 | 0.85 | 73 | -0.17 | 0.18 | 0.31 | 0.397 | 0.81 | 73 |
| **Model fit metrics** | | | | | | | | | | | | |
| **Residual sum of squares (RSS)** | 26.25 | | | | | | 24.54 | | | | | |
| ***AIC*** | 200.46 | | | | | | 187.57 | | | | | |
| ***$AIC_c$*** | 260.01 | | | | | | 247.12 | | | | | |
| $R^2$ | 0.86 | | | | | | 0.87 | | | | | |
| **Adj.** $R^2$ | 0.82 | | | | | | 0.83 | | | | | |

**Table 11. Results for MGWR modelling of mobility reduction, 24 and 31 March.**

| Mobility measured compare to the baseline week | 24 March 2020 | | | | | | 31 March 2020 | | | | | |
|---|---|---|---|---|---|---|---|---|---|---|---|---|
| | Min. | 1st Qu. | Median | 3rd Qu. | Max. | Band width | Min. | 1st Qu. | Median | 3rd Qu. | Max. | Band width |
| **(Intercept)** | -0.04 | 0.08 | 0.09 | 0.216 | 0.29 | 98 | 0.11 | 0.13 | 0.13 | 0.131 | 0.14 | 188 |
| ***Socio-economic status*** | | | | | | | | | | | | |
| Share of households in top household income quintile at national level | 0.16 | 0.17 | 0.43 | 0.672 | 0.75 | 93 | 0.13 | 0.15 | 0.38 | 0.583 | 0.64 | 93 |
| Share of non-English speaker | 0.27 | 0.27 | 0.27 | 0.277 | 0.29 | 188 | 0.27 | 0.28 | 0.28 | 0.298 | 0.31 | 188 |
| Share of social grade C1 (lower middle class) | 0.16 | 0.16 | 0.17 | 0.183 | 0.20 | 188 | 0.15 | 0.17 | 0.18 | 0.307 | 0.36 | 87 |
| ***Accessibility*** | | | | | | | | | | | | |
| Resident population density (1,000 inhabitants per $km^2$) | 0.30 | 0.32 | 1.02 | 1.127 | 1.32 | 67 | 0.28 | 0.51 | 0.85 | 1.104 | 1.57 | 26 |
| ***Activity commitments*** | | | | | | | | | | | | |
| Share of self-employed workers | -0.93 | -0.59 | -0.49 | -0.082 | 0.01 | 46 | -0.90 | -0.67 | -0.63 | -0.092 | -0.06 | 65 |
| ***Population health*** | | | | | | | | | | | | |
| Share of population in poor health | -0.09 | -0.05 | 0.07 | 0.157 | 0.44 | 62 | -0.12 | -0.08 | 0.03 | 0.160 | 0.55 | 55 |
| **Model fit metrics** | | | | | | | | | | | | |
| **Residual sum of squares (RSS)** | 32.62 | | | | | | 26.63 | | | | | |
| $AIC_c$ | 279.82 | | | | | | 269.73 | | | | | |
| $R^2$ | 0.83 | | | | | | 0.86 | | | | | |
| **Adj.** $R^2$ | 0.79 | | | | | | 0.81 | | | | | |

**Table 12. Results for MGWR modelling of mobility reduction, 7 and 14 April.**

| Mobility measured compare to the baseline week | 7 April 2020 | | | | | | 14 April 2020 | | | | | |
|---|---|---|---|---|---|---|---|---|---|---|---|---|
| | Min. | 1st Qu. | Median | 3rd Qu. | Max. | Band width | Min. | 1st Qu. | Median | 3rd Qu. | Max. | Band width |
| **(Intercept)** | 0.23 | 0.24 | 0.25 | 0.251 | 0.27 | 188 | 0.21 | 0.22 | 0.23 | 0.236 | 0.24 | 188 |
| ***Socio-economic status*** | | | | | | | | | | | | |
| Share of households in top household income quintile at national level | 0.15 | 0.28 | 0.57 | 0.843 | 1.06 | 65 | 0.20 | 0.23 | 0.54 | 0.791 | 0.90 | 82 |
| Share of non-English speaker | 0.29 | 0.30 | 0.30 | 0.308 | 0.32 | 188 | 0.28 | 0.28 | 0.29 | 0.300 | 0.31 | 188 |
| Share of social grade C1 (lower middle class) | 0.13 | 0.15 | 0.15 | 0.156 | 0.17 | 188 | 0.08 | 0.14 | 0.17 | 0.243 | 0.35 | 81 |
| ***Accessibility*** | | | | | | | | | | | | |
| Resident population density (1,000 inhabitants per km$^2$) | 0.15 | 0.41 | 0.86 | 1.081 | 1.87 | 24 | 0.20 | 0.39 | 0.92 | 1.108 | 1.51 | 26 |
| ***Activity commitments*** | | | | | | | | | | | | |
| Share of self-employed workers | -0.91 | -0.58 | -0.44 | -0.094 | -0.02 | 46 | -0.92 | -0.64 | -0.50 | -0.093 | 0.02 | 36 |
| ***Population health*** | | | | | | | | | | | | |
| Share of population in poor health | 0.03 | 0.17 | 0.22 | 0.311 | 0.55 | 66 | -0.01 | 0.11 | 0.18 | 0.256 | 0.47 | 65 |
| **Model fit metrics** | | | | | | | | | | | | |
| **Residual sum of squares (RSS)** | 23.80 | | | | | | 22.06 | | | | | |
| $AIC_c$ | 255.94 | | | | | | 247.78 | | | | | |
| $R^2$ | 0.87 | | | | | | 0.88 | | | | | |
| **Adj.** $R^2$ | 0.83 | | | | | | 0.84 | | | | | |

#### Measuring mobility reductions of a 7 day rolling average around each day, compared to the baseline week.

- A comparison between a 7-day rolling average of the median radius of gyration across England around selected Tuesdays and the baseline week (3 March 2020 ± 3 days) mobility levels.
- Table 13 presents the model fit metrics among the developed models using the measured mobility levels by a 7-day rolling average compared to the baseline week.
- Table 14 shows the estimated coefficients of step-wise linear regression models by date.
- Table 15 to Table 18 show the range of estimated coefficients and model fit metrics for the GWR and MGWR models by date.

**Table 13. Assessment of spatial heterogeneity in regression modelling, the dependent variable was estimated by comparing the mobility level on a 7 day rolling average around each of the Tuesdays with baseline week (3 March 2020 ± 3 days) estimates.**

| Modelling approach | Model criterion | 24 March | 31 March | 7 April | 14 April |
|---|---|---|---|---|---|
| **OLS** | Adj. $R^2$ | 0.69 | 0.69 | 0.69 | 0.69 |
| | $AIC$ | 327.14 | 329.92 | 329.33 | 329.92 |
| **GWR** | Adj. $R^2$ | 0.83 | 0.82 | 0.82 | 0.83 |
| | $AIC$ | 190.10 | 196.04 | 190.75 | 188.95 |
| | $AIC_{SLRM} - AIC_{GWR}$ | 137.04 | 133.88 | 138.58 | 140.98 |
| | $AICc$ | 249.65 | 255.59 | 250.30 | 248.49 |
| **MGWR** | Adj. $R^2$ | 0.81 | 0.81 | 0.83 | 0.83 |
| | RSS | 28.38 | 27.21 | 22.03 | 22.89 |
| | $AICc$ | 253.76 | 259.65 | 249.78 | 250.65 |
| | $\Delta AIC_{c_{MGWR}} = (AIC_{c_{GWR}} - AIC_{c_{MGWR}})$ | -4.10 | -4.06 | 0.51 | -2.15 |

Note: GWR and MGWR both uses the bi-square Kernel function and calibrates the bandwidth on the basis of *k*=4 nearest neighbours to generate the local weightings.

**Table 14. Step-wise linear regression model for mobility reduction, by date.**

| Mobility measured using a 7-day rolling average, and compare to the baseline week | 24 March 2020 | | 31 March 2020 | | 7 April 2020 | | 14 April 2020 | |
|---|---|---|---|---|---|---|---|---|
| | Coeff (SD) | LMG | Coeff (SD) | LMG | Coeff (SD) | LMG | Coeff (SD) | LMG |
| **(Intercept)** | 0.00 (0.04) | - | 0.00 (0.04) | - | 0.00 (0.04) | - | 0.000 (0.04) | - |
| ***Socio-economic status*** | | | | | | | | |
| Share of households in top household income quintile at national level | 0.56*** (0.08) | 0.15 | 0.59*** (0.08) | 0.16 | 0.58*** (0.08) | 0.15 | 0.59*** (0.08) | 0.17 |
| Share of non-English speaker | 0.46*** (0.07) | 0.29 | 0.45*** (0.07) | 0.27 | 0.45*** (0.07) | 0.27 | 0.43*** (0.07) | 0.26 |
| Share of social grade C1 (lower middle class) | 0.32*** (0.06) | 0.08 | 0.36*** (0.06) | 0.08 | 0.36*** (0.06) | 0.08 | 0.35*** (0.06) | 0.09 |
| ***Accessibility*** | | | | | | | | |
| Resident population density (1,000 inhabitants per km$^2$) | 0.42*** (0.08) | 0.34 | 0.41*** (0.08) | 0.33 | 0.42*** (0.08) | 0.33 | 0.43*** (0.08) | 0.34 |
| ***Activity commitments*** | | | | | | | | |
| Share of self-employed workers | -0.57*** (0.07) | 0.10 | -0.62*** (0.07) | 0.12 | -0.63*** (0.07) | 0.13 | -0.61*** (0.07) | 0.12 |
| ***Population health*** | | | | | | | | |
| Share of population in bad health | 0.23*** (0.08) | 0.03 | 0.28*** (0.08) | 0.04 | 0.27*** (0.08) | 0.04 | 0.26*** (0.08) | 0.04 |
| **Model fit metrics** | | | | | | | | |
| $R^2$ | | 0.70 | | 0.70 | | 0.70 | | 0.70 |
| Adjusted $R^2$ | | 0.69 | | 0.69 | | 0.69 | | 0.69 |
| Residual Std. Error (df = 184) | | 0.56 | | 0.56 | | 0.56 | | 0.56 |
| F Statistic (df = 6; 184) | | 71.52*** | | 70.04*** | | 70.36*** | | 70.04*** |

**Note: *$p$<0.1; **$p$<0.05; ***$p$<0.01.**

**Table 15. Results for GWR modelling of mobility reduction, 24 and 31 March.**

| Mobility measured using a 7-day rolling average, compare to the baseline week | 24 March 2020 | | | | | | 31 March 2020 | | | | | |
|---|---|---|---|---|---|---|---|---|---|---|---|---|
| | **Min.** | **1st Qu.** | **Median** | **3rd Qu.** | **Max.** | **Band width** | **Min.** | **1st Qu.** | **Median** | **3rd Qu.** | **Max.** | **Band width** |
| **(Intercept)** | -0.17 | 0.10 | 0.23 | 0.350 | 0.55 | 73 | -0.25 | 0.11 | 0.23 | 0.353 | 0.53 | 73 |
| ***Socio-economic status*** | | | | | | | | | | | | |
| Share of households in top household income quintile at national level | 0.20 | 0.35 | 0.605 | 1.05 | 1.38 | 73 | 0.22 | 0.40 | 0.68 | 1.089 | 1.52 | 73 |
| Share of non-English speaker | 0.07 | 0.31 | 0.364 | 0.67 | 0.86 | 73 | 0.02 | 0.32 | 0.39 | 0.710 | 0.94 | 73 |
| Share of social grade C1 (lower middle class) | 0.02 | 0.17 | 0.266 | 0.40 | 0.62 | 73 | 0.08 | 0.21 | 0.33 | 0.462 | 0.70 | 73 |
| ***Accessibility*** | | | | | | | | | | | | |
| Resident population density (1,000 inhabitants per km$^2$) | 0.10 | 0.25 | 0.739 | 1.15 | 1.27 | 73 | 0.11 | 0.22 | 0.66 | 1.123 | 1.28 | 73 |
| ***Activity commitments*** | | | | | | | | | | | | |
| Share of self-employed workers | -1.25 | -0.73 | -0.621 | -0.16 | -0.10 | 73 | -1.30 | -0.79 | -0.69 | -0.210 | -0.11 | 73 |
| ***Population health*** | | | | | | | | | | | | |
| Share of population in poor health | -0.23 | 0.10 | 0.202 | 0.32 | 0.77 | 73 | -0.17 | 0.18 | 0.27 | 0.372 | 0.81 | 73 |
| **Model fit metrics** | | | | | | | | | | | | |
| **Residual sum of squares (RSS)** | | | | | | 24.86 | | | | | | 25.65 |
| ***AIC*** | | | | | | 190.10 | | | | | | 196.04 |
| ***AIC$_c$*** | | | | | | 249.65 | | | | | | 255.59 |
| ***$R^2$*** | | | | | | 0.87 | | | | | | 0.87 |
| **Adj. $R^2$** | | | | | | 0.83 | | | | | | 0.82 |

**Table 16. Results for GWR modelling of mobility reduction, 7 and 14 April.**

| Mobility measured using a 7-day rolling average, compare to the baseline week | 7 April 2020 | | | | | | 14 April 2020 | | | | | |
|---|---|---|---|---|---|---|---|---|---|---|---|---|
| | Min. | 1st Qu. | Median | 3rd Qu. | Max. | Band width | Min. | 1st Qu. | Median | 3rd Qu. | Max. | Band width |
| **(Intercept)** | -0.25 | 0.14 | 0.23 | 0.355 | 0.53 | 73 | -0.21 | 0.16 | 0.25 | 0.349 | 0.50 | 73 |
| ***Socio-economic status*** | | | | | | | | | | | | |
| Share of households in top household income quintile at national level | 0.24 | 0.41 | 0.67 | 1.011 | 1.58 | 73 | 0.25 | 0.43 | 0.68 | 1.068 | 1.55 | 73 |
| Share of non-English speaker | 0.04 | 0.33 | 0.40 | 0.575 | 0.96 | 73 | 0.02 | 0.32 | 0.39 | 0.570 | 0.90 | 73 |
| Share of social grade C1 (lower middle class) | 0.09 | 0.22 | 0.33 | 0.443 | 0.64 | 73 | 0.06 | 0.20 | 0.30 | 0.435 | 0.65 | 73 |
| ***Accessibility*** | | | | | | | | | | | | |
| Resident population density (1,000 inhabitants per km$^2$) | 0.14 | 0.22 | 0.62 | 1.171 | 1.35 | 73 | 0.12 | 0.20 | 0.63 | 1.139 | 1.31 | 73 |
| ***Activity commitments*** | | | | | | | | | | | | |
| Share of self-employed workers | -1.36 | -0.79 | -0.68 | -0.216 | -0.12 | 73 | -1.32 | -0.78 | -0.66 | -0.212 | -0.11 | 73 |
| ***Population health*** | | | | | | | | | | | | |
| Share of population in poor health | -0.18 | 0.18 | 0.27 | 0.320 | 0.79 | 73 | -0.19 | 0.18 | 0.29 | 0.358 | 0.82 | 73 |
| **Model fit metrics** | | | | | | | | | | | | |
| Residual sum of squares (RSS) | 24.95 | | | | | | 24.71 | | | | | |
| *AIC* | 190.75 | | | | | | 188.95 | | | | | |
| $AIC_c$ | 250.30 | | | | | | 248.49 | | | | | |
| $R^2$ | 0.87 | | | | | | 0.87 | | | | | |
| Adj. $R^2$ | 0.82 | | | | | | 0.83 | | | | | |

**Table 17. Results for MGWR modelling of mobility reduction, 24 and 31 March.**

| Mobility measured using a 7-day rolling average, compare to the baseline week | 24 March 2020 | | | | | | 31 March 2020 | | | | | |
|---|---|---|---|---|---|---|---|---|---|---|---|---|
| | **Min.** | **1st Qu.** | **Median** | **3rd Qu.** | **Max.** | **Band width** | **Min.** | **1st Qu.** | **Median** | **3rd Qu.** | **Max.** | **Band width** |
| **(Intercept)** | 0.08 | 0.10 | 0.10 | 0.110 | 0.12 | 188 | 0.13 | 0.14 | 0.15 | 0.150 | 0.16 | 188 |
| ***Socio-economic status*** | | | | | | | | | | | | |
| Share of households in top household income quintile at national level | 0.16 | 0.18 | 0.41 | 0.601 | 0.68 | 93 | 0.18 | 0.20 | 0.44 | 0.647 | 0.73 | 96 |
| Share of non-English speaker | 0.30 | 0.31 | 0.31 | 0.309 | 0.32 | 188 | 0.29 | 0.29 | 0.29 | 0.297 | 0.31 | 188 |
| Share of social grade C1 (lower middle class) | 0.14 | 0.14 | 0.15 | 0.178 | 0.20 | 184 | 0.17 | 0.17 | 0.18 | 0.209 | 0.23 | 184 |
| ***Accessibility*** | | | | | | | | | | | | |
| Resident population density (1,000 inhabitants per $km^2$) | 0.28 | 0.30 | 0.88 | 1.005 | 1.25 | 55 | 0.25 | 0.35 | 0.81 | 1.054 | 1.42 | 35 |
| ***Activity commitments*** | | | | | | | | | | | | |
| Share of self-employed workers | -0.90 | -0.61 | -0.50 | -0.076 | 0.01 | 46 | -0.91 | -0.61 | -0.51 | -0.097 | 0.00 | 46 |
| ***Population health*** | | | | | | | | | | | | |
| Share of population in poor health | -0.14 | -0.04 | 0.05 | 0.154 | 0.45 | 62 | -0.06 | 0.03 | 0.11 | 0.222 | 0.51 | 62 |
| **Model fit metrics** | | | | | | | | | | | | |
| Residual sum of squares (RSS) | | | | | | 28.38 | | | | | | 27.21 |
| $AIC_c$ | | | | | | 253.76 | | | | | | 259.65 |
| $R^2$ | | | | | | 0.85 | | | | | | 0.86 |
| Adj. $R^2$ | | | | | | 0.81 | | | | | | 0.81 |

**Table 18. Results for MGWR modelling of mobility reduction, 7 and 14 April.**

| Mobility measured using a 7-day rolling average, compare to the baseline week | 7 April 2020 | | | | | | 14 April 2020 | | | | | |
|---|---|---|---|---|---|---|---|---|---|---|---|---|
| | **Min.** | **1st Qu.** | **Median** | **3rd Qu.** | **Max.** | **Band width** | **Min.** | **1st Qu.** | **Median** | **3rd Qu.** | **Max.** | **Band width** |
| **(Intercept)** | 0.21 | 0.23 | 0.23 | 0.236 | 0.25 | 188 | 0.17 | 0.19 | 0.20 | 0.202 | 0.21 | 188 |
| ***Socio-economic status*** | | | | | | | | | | | | |
| Share of households in top household income quintile at national level | 0.14 | 0.26 | 0.54 | 0.784 | 0.96 | 65 | 0.16 | 0.18 | 0.45 | 0.719 | 0.83 | 93 |
| Share of non-English speaker | 0.30 | 0.30 | 0.31 | 0.315 | 0.33 | 188 | 0.26 | 0.27 | 0.27 | 0.281 | 0.29 | 188 |
| Share of social grade C1 (lower middle class) | 0.18 | 0.18 | 0.19 | 0.210 | 0.23 | 185 | 0.13 | 0.16 | 0.18 | 0.268 | 0.36 | 84 |
| ***Accessibility*** | | | | | | | | | | | | |
| Resident population density (1,000 inhabitants per km$^2$) | 0.18 | 0.45 | 0.87 | 1.081 | 1.70 | 24 | 0.24 | 0.41 | 0.91 | 1.118 | 1.54 | 28 |
| ***Activity commitments*** | | | | | | | | | | | | |
| Share of self-employed workers | -1.02 | -0.64 | -0.50 | -0.128 | 0.00 | 36 | -0.99 | -0.66 | -0.54 | -0.101 | 0.02 | 36 |
| ***Population health*** | | | | | | | | | | | | |
| Share of population in poor health | -0.01 | 0.16 | 0.21 | 0.278 | 0.51 | 65 | -0.06 | 0.03 | 0.11 | 0.210 | 0.46 | 62 |
| **Model fit metrics** | | | | | | | | | | | | |
| Residual sum of squares (RSS) | 22.03 | | | | | | 22.89 | | | | | |
| $AIC_c$ | 249.78 | | | | | | 250.65 | | | | | |
| $R^2$ | 0.88 | | | | | | 0.88 | | | | | |
| Adj. $R^2$ | 0.83 | | | | | | 0.83 | | | | | |
